# Global, regional, and national burden of multiple sclerosis from 1990 to 2021: A systematic analysis for the Global Burden of Disease Study 2021

**DOI:** 10.1101/2025.04.17.25325986

**Authors:** Wei She, Ruilin Li, Yuping Zou, Linna Yuan, Yingnan Ma, Haiyan Chen, Siyu Wei, Jiacheng Wang, Junxian Tao, Chen Sun, Shuo Bi, Xuying Guo, Hongsheng Tian, Jing Xu, Zhenwei Shang, Hongchao Lv, Yongshuai Jiang, Mingming Zhang

**Affiliations:** College of Bioinformatics Science and Technology, Harbin Medical University, Harbin, China

**Author notes:** Correspondence: Mingming Zhang, College of Bioinformatics Science and Technology, Harbin Medical University, 194 Xuefu Road, Nangang District, Harbin, Heilongjiang Province, China.;, Yongshuai Jiang, College of Bioinformatics Science and Technology, Harbin Medical University, 194 Xuefu Road, Nangang District, Harbin, Heilongjiang Province, China.;, Hongchao Lv, College of Bioinformatics Science and Technology, Harbin Medical University, 194 Xuefu Road, Nangang District, Harbin, Heilongjiang Province, China. These authors contributed equally to this work (Wei She, Ruilin Li, Yuping Zou, Linna Yuan, Yingnan Ma).

**Keywords:** Multiple sclerosis, Global burden of disease, Risk factor

## Abstract

**Background:** Multiple sclerosis (MS) is an autoimmune disease resulting from the immune system mistakenly attacks the brain and spinal cord. In recent decades, the prevalence of MS has increased, leading to a significant global health burden.

**Methods:** The study presented a comprehensive analysis of the incidence, prevalence, deaths, and disability-adjusted life years (DALYs) in MS using data from the Global Burden of Diseases, Injuries, and Risk Factors Study (GBD) 2021. Subsequently, Mendelian randomization (MR) was employed to investigate potential risk factors associated with MS. Finally, the Bayesian age-period-cohort (BAPC) model was used to predict the future trend of MS.

**Results:** From 1990 to 2021, the cases of MS increased from 1,004,659 to 1,887,767, while the age-standardized rate (ASR) remained relatively stable. Areas with higher Socio-demographic Index (SDI), such as Western Europe and High-income North America, exhibited a greater disease burden, whereas areas with lower SDI exhibited a higher ASR growth. The highest incidence rate was observed in individuals aged 30 to 34 years. The number of cases in females was approximately twice that in males. In addition to smoking, other risk factors, including body mass index, waist circumference, high-density lipoprotein cholesterol, and overweight, were identified by MR. It is anticipated that the ASIR of MS will continue to remain at a relatively high level until 2050.

**Conclusions:** The burden of MS was increasing globally, with variations observed across different geographies, sexes, and age groups, presenting a significant challenge to public health. This study provided a comprehensive investigation into the burden of MS, which would assist in the allocation of resources and the development of targeted prevention and control strategies.

## Introduction

Multiple sclerosis (MS) is an autoimmune disease characterized by the inflammatory demyelination of the central nervous system, resulting from a complex interplay between environmental and genetic factors.^1,2^ The diagnosis of MS is based on the McDonald criteria, which is considered the gold standard and integrates clinical evaluation, magnetic resonance imaging, and cerebrospinal fluid analysis to confirm evidence of dissemination in space and time.^3^ MS is one of the most common causes of neurological disability in the young adult population, with an average age of diagnosis being 32 years old.^4^ Compared to the healthy population, MS patients had a reduced life expectancy of 7 years and their mortality rate is nearly three times.^5^ Furthermore, MS has a significant economic impact globally. The annual economic cost of MS in the USA was estimated at US$85.4 billion, and the cost of treating MS patients exceeds the nominal GDP per capita of some countries in the Middle East.^6,7^ Therefore, an analysis of the global MS disease burden is imperative.

The Global Burden of Diseases, Injuries, and Risk Factors Study (GBD) 2021 is a valuable tool for investigating the burden of disease. Currently, several studies based on the GBD have reported the burden of MS. Zhang et al. exclusively examined the burden changes of MS across 33 province-level administrative units in China, without considering other regions.^8^ Additionally, Qian Z et al. reported the global, national, and regional burden of MS, but they did not provide trends in prevalence or detailed analysis of risk factors.^9^ Other studies had examined the global disease burden of autoimmune diseases, but had not focused on a specific analysis of MS.^10-12^ The GBD Neurology Collaborators performed a comprehensive analysis of the global burden of MS, but the data presented was outdated and limited insights into the current disease burden.^13^

With the increase in MS-related public health data, it has become feasible to conduct a systematic, detailed, and up-to-date analysis of the MS burden. Consequently, our study analyzed the current disease burden at the global, regional, and national levels, as well as the differences by sex and age. Furthermore, we also discussed the risk factors and future trends in order to provide insights into MS prevention.

## Methods Data

The GBD database (http://ghdx.healthdata.org/gbd-results-tool) is a developed and shared data platform to quantify health loss across various locations and over time.^14^ We performed a secondary comprehensive analysis of the data on incidence, prevalence, deaths, years of life lost (YLL), years lived with disability (YLD), and disability-adjusted life years (DALYs) of MS across 204 countries and territories provided by GBD 2021. In GBD 2021, the incidence and prevalence mainly referred to McDonald’s diagnostic criteria and were modeled using DisMod-MR 2.1 (Disease Modeling Meta Regression; version 2.1). MS’s deaths predicted using Cause of Death Ensemble modeling (CODEm), and GBD 2021 evaluated cause-specific death rates using CODEm. YLDs were calculated through a microsimulation process, with adjusted through consequences of MS and disability weights for each sequela. YLLs were computed by multiplying the number of estimated deaths by the predicted life expectancy by age, sex, location, and year.(Detailed data selection in appendix p2) In addition, the genome-wide association study(GWAS) summary data used to explore the relationship between MS and risk factors comes from the Integrative Epidemiology Unit open GWAS project (Open GWAS, https://gwas.mrcieu.ac.uk) study. (For details, please refer to appendix p4)

### Socio-demographic Index (SDI)

SDI is a comprehensive indicator of the development status of a region derived from lag-distributed income per capita, mean years of education attainment, and the total fertility rate under 25 years.^15^ In the GBD study, countries and territories were classified into five levels (low SDI, low-middle SDI, middle SDI, high-middle SDI, and high SDI) based on SDI quintiles (appendix p2).^16^

### Age-standardized rate (ASR)

Age is an important factor affecting disease burden. The ASR can be obtained through the adjustment of the age distribution by applying the age-specific rates of each population to a standard population. The GBD 2021 calculated ASR per 100,000 population using a unified GBD standard population structure, which offers a reasonable index for comparing disease burden across different regions and countries.^17^ (Detailed data selection in appendix p2)

### Average annual percentage change (AAPC)

The AAPC, calculated by the joinpoint regression in joinpoint software, can evaluate development trends of age-standardized incidence rate (ASIR), age-standardized prevalence rate (ASPR), age-standardized mortality rate (ASMR), and age-standardized DALY rate (ASDR) over time.^18,19^

### Two-sample Mendelian randomization (MR)

MR can investigate disease risk factors and evaluate the causal relationships between risk factors and diseases. We selected Single Nucleotide Polymorphism (SNP) strongly associated with risk factors as instrumental variables(IVs). To ensure the validity of these IVs, we removed linkage disequilibrium (LD) between SNPs, thereby minimizing confounding effects and improving the independence of each SNP. The MR analysis was performed using the R package “TwoSampleMR (version 0.6.1)”.^20^ (Detailed information can be found in the appendix p3)

### Bayesian age-period-cohort (BAPC) analysis

The BAPC model was employed to predict the future burden of disease by analyzing the impact of three types of time-related variations – age, period, and cohort – on the incidence of MS. The model is capable of utilizing both sample and prior information to obtain unique parameter estimates, thereby ensuring robust and reliable results.^21,22^ In this study, BAPC analysis was conducted using the “BAPC” package (version 0.0.36) and the “INLA” package (version 24.2.9) in the R language.^23^

## Results

### The global burden of MS from 1990 to 2021

In 1990, the global prevalence of MS cases was 1,004,659 (95% uncertainty interval (UI): 868,374 to 1,165,224). By 2021, this number had increased to 1,887,767 (95% UI: 1,688,654 to 2,113,707), with an increase of 87.901% (95% UI: 80.847% to 95.534%) compared to 1990. The global ASPR decreased from 22.26 (95% UI: 19.30 to 25.65) per 100,000 to 22.17 (95% UI: 19.77 to 24.82) per 100,000, with an AAPC of -0.0108 (95% confidence interval (CI): -0.0348 to -0.0132) from 1990 to 2021 (Table S1). Despite the global ASPR of MS exhibited a modest decline, there were 16,302 (95% UI: 15,357 to 17,039) deaths due to MS in 2021 (Table S1, appendix p3, p4).

### The regional burden of MS from 1990 to 2021

At the regional level, Western Europe had the highest prevalence of MS cases with 544,964 (95% UI: 485,454 to 613,732) in 2021. Additionally, Western Europe ranked second with the ASPR of 91.36 (95% UI: 80.90 to 102.99), while the highest ASPR was observed in High-income North America (103.61, 95% UI: 96.44 to 111.33). Regarding deaths, the numbers had increased in all regions except Central Europe (from 1,301 (95% UI: 1,233 to 1,383) to 1,041 (95% UI: 946 to 1,146)) and Eastern Europe (from 1,393 (95% UI: 1,341 to 1,451) to 997 (95% UI: 889 to 1,110)). The most notable increase in ASMR was observed in Central Latin America (AAPC: 2.0793) which also exhibited the greatest increases in ASPR (AAPC: 1.4686), ASIR (AAPC: 1.0944), and ASDR (AAPC: 2.0793) (Table S1, appendix p4, p5).

### The national burden of MS from 1990 to 2021

In 2021, only nine countries globally reported fewer cases than in 1990, which were predominantly located at a distance from the equator (Figure S1). In contrast, many other countries exhibited a notable increase in the number of MS cases, with Qatar (1384.979%, 95% UI: 1184.123% to 1626.707%) experiencing the most pronounced increment. The ASPR of MS ranged from 1.45 to 161.60 cases per 100,000 population of 204 countries and territories in 2021. Sweden (161.60, 95% UI: 140.22 to 186.95), Canada (134.20, 95% UI: 130.83 to 137.70), and Norway (131.53, 95% UI: 111.04 to 154.17) had the three highest ASPR (Figure 1). The United Kingdom had the highest ASMR (1.34, 95% UI: 1.27 to 1.40) and ASDR (71.30, 95% UI: 61.83 to 80.79). Compared with 1990, the number of DALYs in 2021 increased in 188 out of 204 countries and territories, with the largest increase still in Qatar (1,380.277%, 95% UI: 1,094.6% to 1,778.844%) (appendix p6, p7).

**Figure 1.**
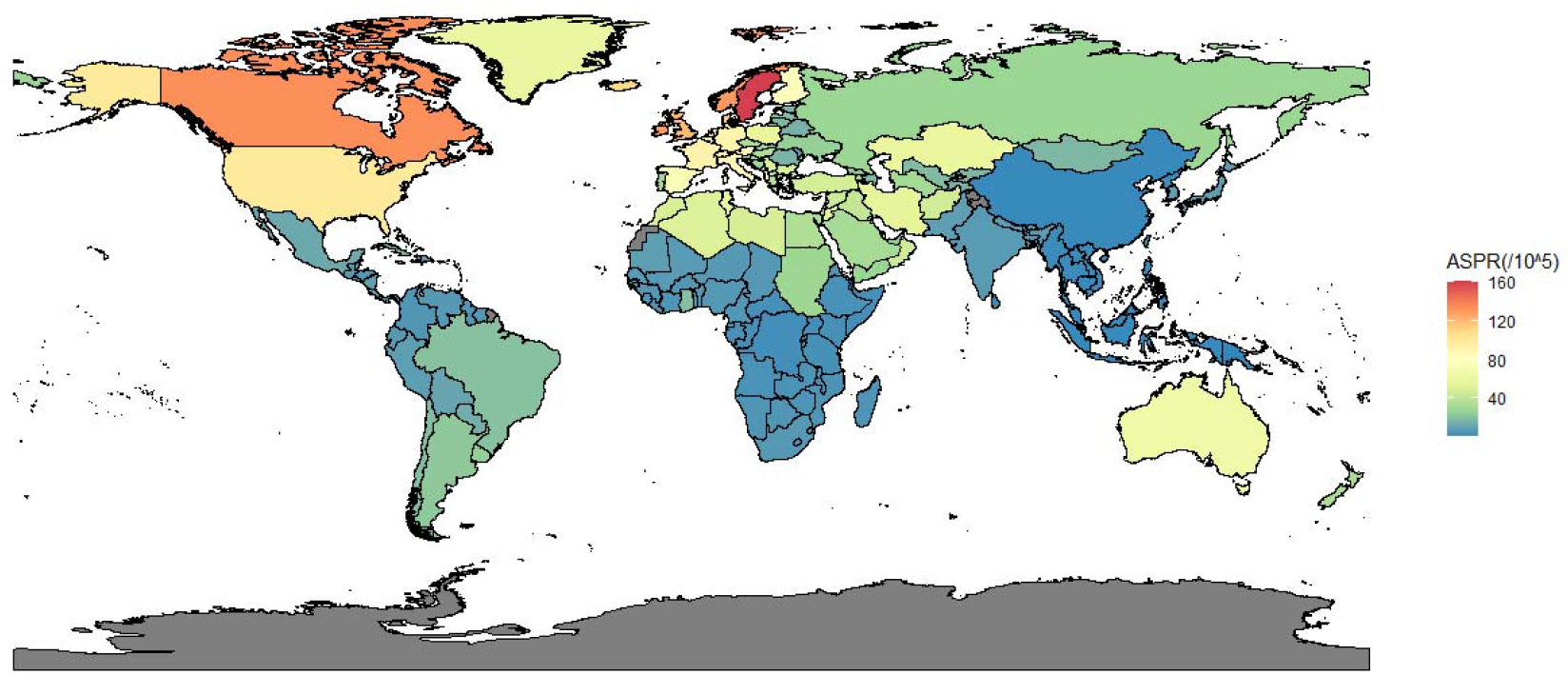
World map of ASPR per 100,000 population distribution for global MS in 2021. The black areas represent areas where no or few people live.

### The impact of age and sex on burden of MS

In 2021, our findings indicated a significantly higher burden of MS in females compared to males, with 1,270,821 cases in females as well as 616,946 in males. The number of prevalence, incidence, deaths, and DALYs consistently increased with advancing age, followed by a decline once the peak was reached (Figure 2). The age group with the highest ASIR was concentrated in the 30-34 years age group. In contrast, the age groups with the highest ASPR and ASDR were in the 60-64 years age group, and the highest ASMR was observed in the 90-94 years age group. Furthermore, the ASPR stabilized at approximately 30 per 100,000 for males and 65 per 100,000 for females following the peak, whereas the ASIR, ASDR and ASMR exhibited a decline.

**Figure 2.**
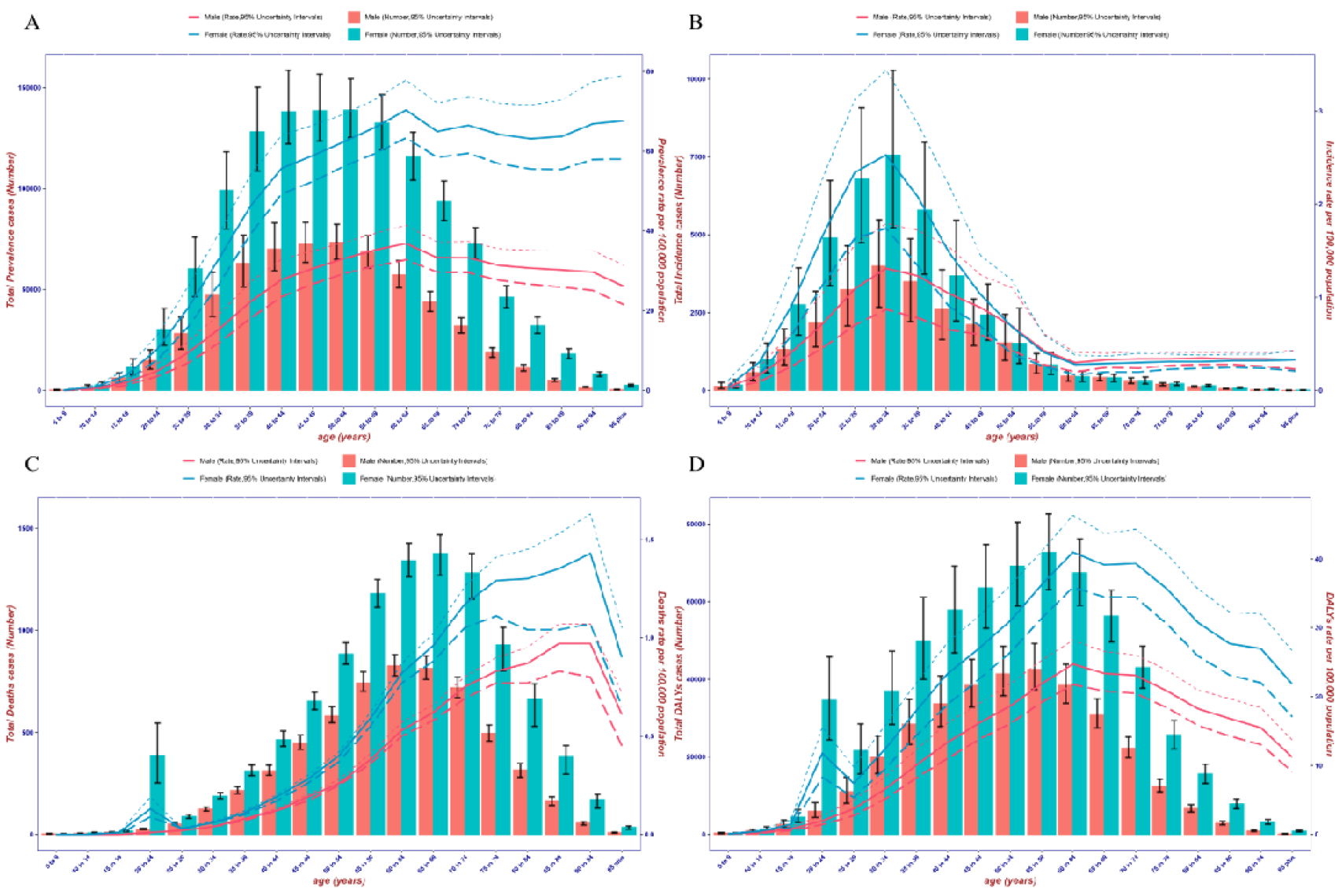
Change trend of the ASR per 100,000 population and number of cases of MS by sexes with age in 2021. (A):ASPR and prevalence cases; (B):ASIR and incidence cases; (C):ASMR and death cases; (D):ASDR and DALY cases; dashed lines indicate 95% upper and lower uncertainty intervals, respectively;the bar chart reflects cases, while the line chart reflects ASR.

### The impact of SDI on burden of MS

From 1990 to 2021, SDI had a significant global impact on ASRs in both males and females. In 2021, the highest ASIR, ASPR, ASDR, and ASMR were observed in high SDI region. Conversely, the ASRs in middle SDI, low-middle SDI, and low SDI regions were significantly lower than those in the other two regions. The ASRs in the high-middle SDI region were largely consistent with that observed globally, and it was the only region to experience a decline in ASRs compared to the other SDI regions. Furthermore, the ASR growth rates in the middle SDI, low-middle SDI, and low SDI regions were significantly higher than in other regions. From a sex perspective, females exhibited higher ASRs than males across all five SDI regions. The difference in MS burden trends between males and females was observed in the high SDI region, where ASDR and ASMR exhibited a decrease for males and an increase for females (Figure 3).

**Figure 3.**
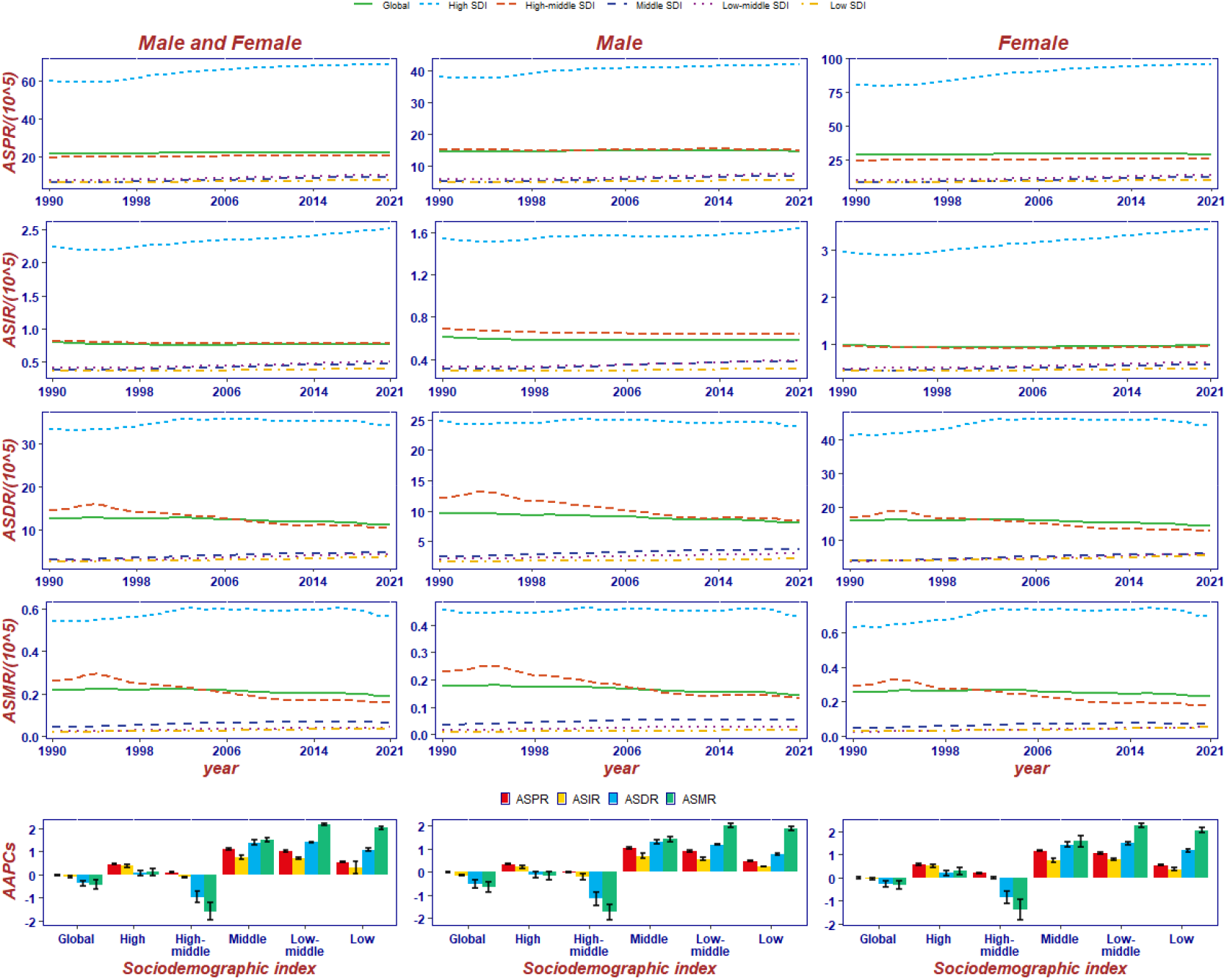
Temporal trends of the ASR per 100,000 population and AAPC of MS in different sex and different SDI regions in global from 1990 to 2021.

Concurrently, our findings indicated that as SDI increased, ASRs demonstrated an accelerating upward trend, suggesting that regions with higher SDI also had higher MS burden in 2021 (Figure 4A-D). The AAPC of ASIR and ASPR exhibited a positive correlation with SDI, as well as the AAPC of ASDR and ASMR exhibited inverted U-shaped curve with SDI (Figure 4E-H). It is noteworthy that the correlation curves of the AAPC and SDI were all above 0, indicating an upward trend in ASRs. The ASPR and ASIR exhibited greater growth in high SDI region, while the ASMR and ASDR demonstrated greater growth in low-middle SDI region (Figure 4E-H).

**Figure 4.**
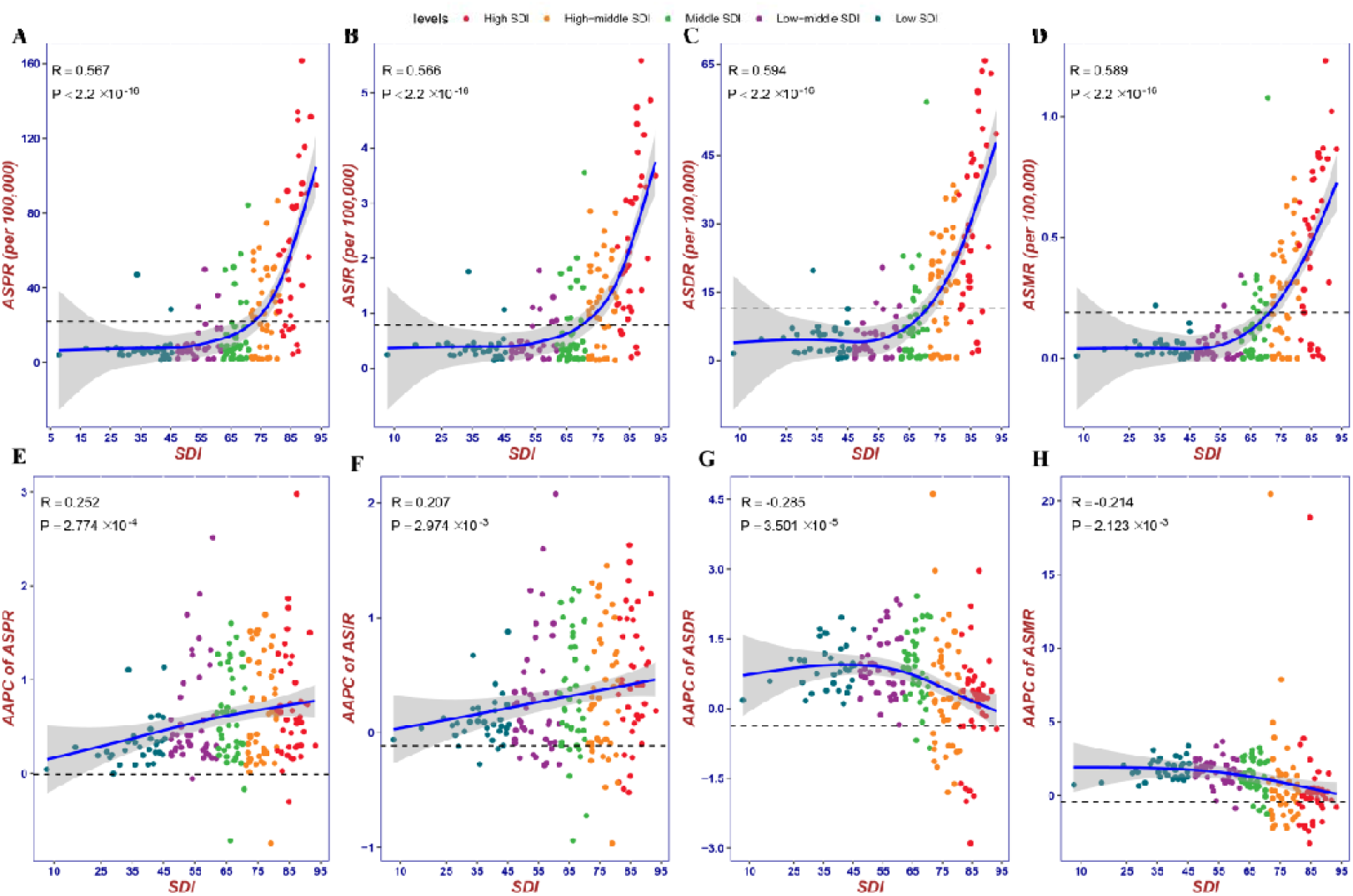
Correlation between SDI and ASR per 100,000 population of MS in 2021, as well as SDI and AAPC of ASR from 1990 to 2021. (A):ASPR; (B):ASIR; (C):ASMR; (D):ASDR; (E):AAPC of ASPR; (F):AAPC of ASIR; (G):AAPC of ASMR; (H):AAPC of ASDR; The dotted line represents the global level.

### The impact of risk factors on burden of MS

In the GBD 2021 study, smoking was reported as the sole risk factor for MS. From 1990 to 2021, the global DALYs percentage of MS caused by smoking decreased from 18.5% to 11.2%. This decrease was observed in almost all regions, with the highest decrease in the high SDI region (from 21.8% to 13.4%). Eastern Europe was the only region where the percentage increased from 13.5% to 14.1% (Figure 5A). The impact of risk factors on the percentage of DALYs exhibited significant variation by sex, and the percentage of DALYs attributed to smoking was higher in males than in females across all regions from 1990 to 2021 (Figure 5B). The deaths attributed to MS due to smoking exhibited a similar trend (appendix p7).

**Figure 5.**
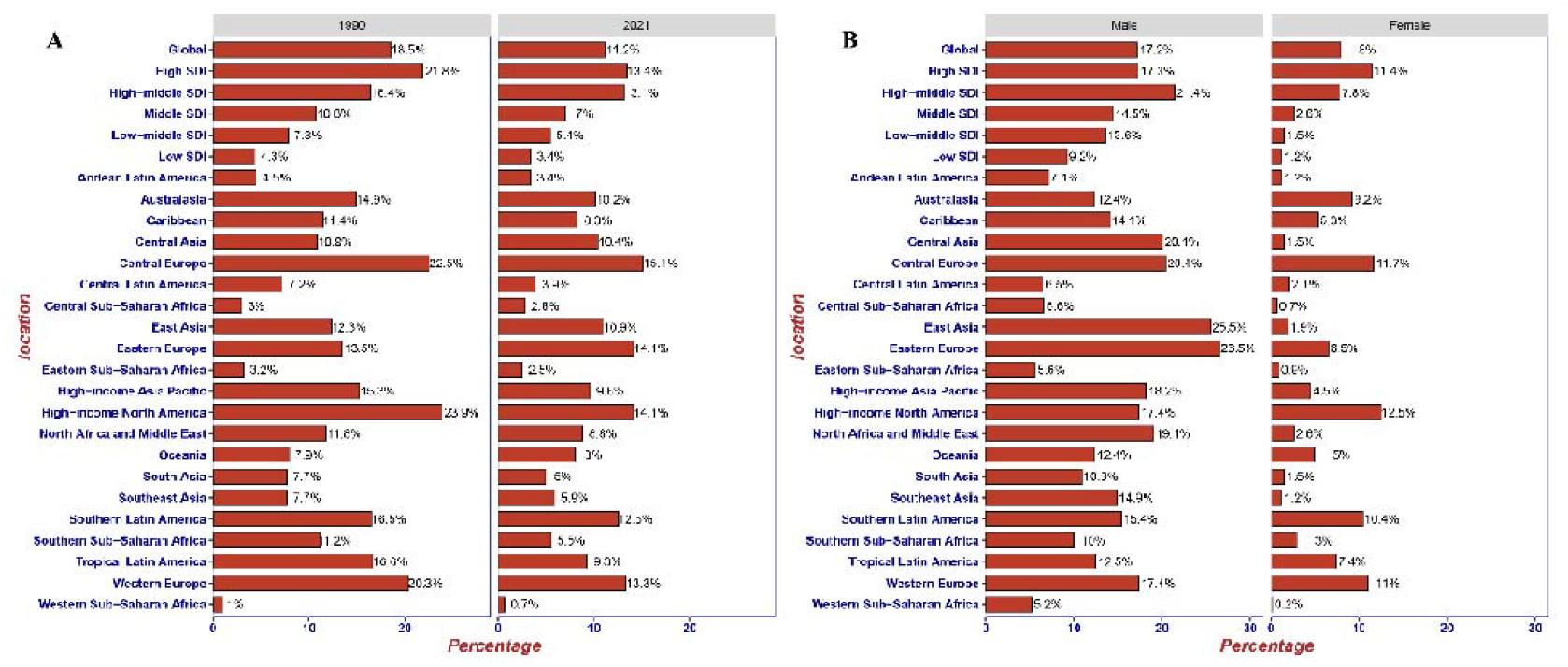
Percentage of MS DALYs caused by smoking in different regions in 1990 and 2021 (A), as well as by sexes(B).

The two-sample MR analysis of the 226 factors in the OpenGWAS database revealed that four factors were causally associated with MS (Figure 6). All of these factors, including body mass index (OR: 1.26, 95% CI: 1.08 to 1.45), waist circumference (OR: 1.35, 95% CI: 1.12 to 1.59), high density lipoprotein cholesterol (OR: 1.13, 95% CI: 1.04 to 1.23), and overweight (OR: 1.20, 95% CI: 1.08 to 1.32), demonstrated a positive association with MS across all methods. This indicated that these factors increased the risk of developing the disease.

**Figure 6.**
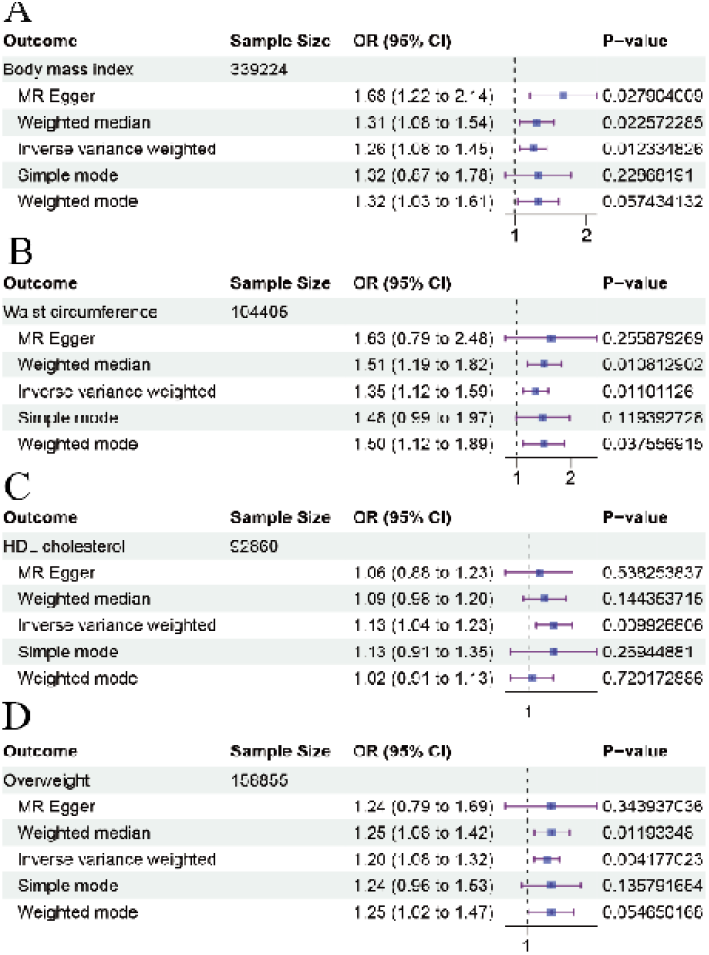
MR forest plot of risk factor with MS. (A) :BMI; (B): waist circumference; (C): HDL-C; (D): overweight; MR: Mendelian randomization; OR = odds ratio.

### ASIR forecasted by BAPC from 2022 to 2050

The BAPC analysis indicated that there will be no significant change in ASIR for males and females until 2050 (Figure 7). The analysis of ASIR across different age groups for both males and females revealed a consistent trend. It was anticipated that, from 2022 to 2050, the female population would continue to exhibit a higher ASIR than the male population. By 2050, the disparity in the number of cases between males and females will be projected to narrow, with 27,781 new cases in males and 48,078 new cases in females. The age of onset in males and females will remain unchanged, reaching they peak at the age of 30-34. (Figure S13, Figure S14).

**Figure 7.**
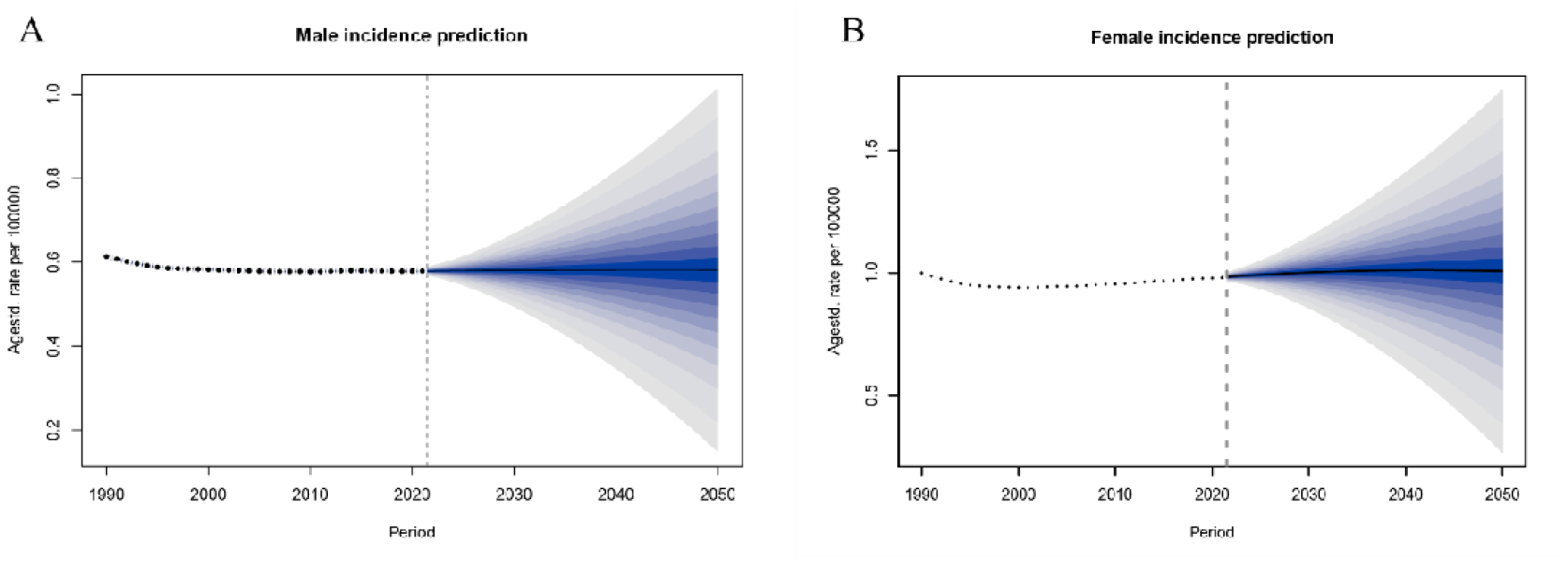
BAPC’s prediction of changes in ASIR per 100,000 population of global MS male (A) and female (B) until 2050. The blue area is uncertain interval.

## Discussion

In this study, we conducted a comprehensive survey on the global, regional, and national burden of MS. From 1990 to 2021, this study revealed that the incidence, prevalence, deaths, and DALYs of MS cases have increased globally, while the ASIR, ASPR, ASMR, and ASDR have decreased. In 2021, there were 1,887,767 prevalent cases, 62,920 incidence cases, 16,302 deaths, and 973,298 DALYs globally(Table S1). As with previous studies, our studies found that high-income North America and Western Europe had the highest burden of MS, while regions such as Southeast Asia and Africa had a lower prevalence of MS.^9^ The burden of MS in Sweden was the most severe in our study, with other studies have shown that the production losses caused by MS in Sweden result in costs greater than medical consumption.^24^ It is important to note that Qatar has experienced significant growth in both the number of cases and the ASR over the past few decades.

We found significant latitude differences in the burden of MS between North America and Western Europe on the ASPR world MS burden distribution map (Figure 1). We extracted the ASPR data of these two regions and compared it with the latitude of the countries (Table S6). Afterwards, we found that the burden of MS in North America and Western Europe decreased from north to south, which is consistent with previous studies.^25,26^ Previous studies had shown that there was a strong latitudinal variation in the burden of MS and the high latitude countries and territories had the higher MS burden.^13,27^ But the influence of latitude appears to be changing. In our study, the ASPR of the high latitude country Greenland was relatively low.

Previous studies have reported a positive correlation between ASDR and SDI, and concluded that the disease burden is more severe in areas with high SDI than in areas with low SDI.^9^ In agreement with these findings, our results also found that ASIR, ASPR, and ASMR had similar correlations with SDI, which confirming that the burden of MS was higher in developed areas than in developing countries.^28^ One reason might be that the overuse of vaccines and antibiotics in developed countries have led to inappropriate changes in the immune system, making people more susceptible to autoimmune diseases.^29^

The prevalence of MS in adulthood varies greatly between males and females. Previous studies have found that the incidence of MS in females increased significantly in the late 20th century.^30,31^ Our study found that the incidence, prevalence, mortality, and DALYs of MS were higher in females across all regions from 1990 to 2021, with the number of cases in females being about twice that of male. The possible causes of this difference may be overuse of gonadal hormones and oral contraceptives, changes in lifestyle and lactation patterns, reduced physical activity, as well as increased stress.^13,32^ This suggestes that we should invest more effort in female to reduce the MS disease burden.

The cause of MS is multifactorial, with environmental and genetic factors affecting the risk of disease, and even different populations of disease risk is different.^25,26^ Currently, the only confirmed risk factor for MS identified in the GBD was smoking. We also found four other risk factors (body mass index, waist circumference, high density lipoprotein cholesterol, and overweight) associated with MS using MR. It is notable that body mass index, waist circumference, and overweight were all measures of obesity, suggesting a possible correlation between them. After conducting principal component analysis on them, we found the contribution for first principal component of body mass index, waist circumference, and overweight, accounting for 97.693%, demonstrating their strong interrelationship.(appendix P9) This indicates that obesity may increase the risk of MS, which is consistent with previous studies.^33^Furthermore, the strongest known risk factor for MS was EBV infection, which was not included in the GBD 2021 study.^34,35^ Vaccines, stress, traumatic events, and allergies may have an impact on MS, but they have not been identified as risk factors.^32^ Therefore, it is necessary to study the etiology of MS in the further.

Like other estimation efforts, our research has its limitations. Firstly, GBD research relies on various assumptions and modeling calculations, which may introduce some uncertainty into our study.^17^ Secondly, due to limited medical services and inadequate disease diagnosis in underdeveloped regions, the true burden in these regions may be underestimated. Additionally, the COVID-19 pandemic has fundamentally altered global health and mortality patterns.^36^ It may have impacted the mortality and disability data of MS, leading us to potentially overestimate the contribution of smoking. Therefore, our results should be viewed as the most likely estimates based on current evidence.

The results of the BAPC showed that the incidence of MS in the future will not change significantly, indicating that the global burden of MS will still considerable in the future. Our findings provided a comprehensive and contemporary understanding of the global MS disease burden, which would facilitate the allocation of resources and the planning of healthcare for the growing number of individuals with MS worldwide.

## Conclusion

MS is a global health issue that had a steady increase in cases over the past 30 years, particularly in developed regions. The disease burden of MS exhibits geographical disparities, with a higher ASR burden observed in regions situated at far from the equator. Additionally, the disease burden was higher in females than in males, with the peak age of incidence being 30-34 years. The causal relationships between MS and body mass index, waist circumference, high-density lipoprotein cholesterol, and overweight were identified through MR. The incidence of MS will be remained relatively stable until 2050, indicating the need for continued attention to this disease. Therefore, prevention and treatment strategies should be focused on young adults, females, and regions with a high disease burden.

## Supporting information

Supplementation of methods and results

## Data Availability

Original data generated and analyzed during this study are included in this published article or in the data repositories listed in References.

https://vizhub.healthdata.org/gbd-results/

## Human Ethics and Consent to Participate declarations

Not applicable.

## Contributors

Yongshuai Jiang, Mingming Zhang and Hongchao Lv designed this study. Wei She, Yuping Zou, Ruilin Li, Yingnan Ma, and Linna Yuan prepared the initial draft and finalised the manuscript with comments from all other authors. Junxian Tao, Jiacheng Wang, and Chen Sun collected and analysed the data. Shuo Bi, Hongsheng Tian, and Jing Xu validated the data. Xuying Guo, Siyu Wei, and Haiyan Chen made important revisions to the manuscript. Zhenwei Shang, Yu Dong and Jingxuan Kang participated in the interpretation of the data and provided important comments on the manuscript. All authors had full access to all the data in the study and were responsible for the integrity of the data and the accuracy of the data analysis. All authors reviewed the draft for criticality and approved the final version.

## Acknowledgments

Thanks to the GBD 2021 database for providing data. This work was supported by the National Natural Science Foundation of China [Grant Nos. 31970651, 92046018]; Program for Young Talents of Basic Research in Universities of Heilongjiang Province [Grant No.YQJH2023036]; Marshal Initiative Funding [Grant No. HMUMIF-22010]; Mathematical Tianyuan Fund of the National Natural Science Foundation of China [Grant No. 12026414].

