## Supplementation of methods and results for "Global, regional, and national burden of multiple sclerosis from 1990 to 2021: A systematic analysis for the Global Burden of Disease Study 2021"

### Method

#### Data sources

The GBD database (http://ghdx.healthdata.org/gbd-results-tool) database is a comprehensive database organized by global health research institutions to assess and analyze the health impact of diseases, injuries and risk factors globally and regionally. The database is led by the Institute for Health Metrics and Evaluation (IHME) at the University of Washington. The main objective of the database is to collect, collate and analyze data on the global burden of various diseases and injuries, including morbidity, mortality, Disability-Adjusted Life Years (DALYs) and other indicators. The GBD database covers 204 countries and territories around the world, with a time range from 1990 to 2021, as well as some forecast years in the future. It not only includes traditional infectious diseases, chronic diseases and other diseases, but also includes mental health problems, injuries, environmental pollution and other factors on health. Through this data, GBD projects are able to assess global health status, identify health trends, and assess the effectiveness of public health policies and interventions.

MS, as a chronic, degenerative, and progressive neurological disease, has also received significant attention in GBD2021. The McDonald diagnostic criteria are clinical, radiological, and laboratory standards used to diagnose multiple sclerosis, and are currently the gold standard for MS diagnosis. The McDonald diagnostic criteria were introduced in 2001 and have undergone multiple revisions.^1^ In GBD2021, the 2017 McDonald diagnostic criteria, Other published criteria (such as Poser, Schumacher, or McAllen criteria) and clinical neurological exam as references for MS, The ICD-10 code for MS is G35. In GBD 2021, the data for MS comes from two sources: one is the systematic review of published studies, and the second is the claim data obtained by the GBD clinical informatics team. New data added in GBD 2021 included Polish claims (2017–2018), and an additional year of USA claims (2017). For the systematic review of published studies data, gender processing is performed based on existing gender stratification or MR-BRT (meta regression Bayesian, regularized, trimmed) models, referencing the global age pattern of the 2017 GBD age specific model for age processing, and referencing the nonfatal database of CODEm (Cause of Death Ensemble model). The CODEm is also the cause of death estimation model for most diseases and injuries in GBD2021, and the death related data used in our article also comes from this model. Then, combined with the claim data, Disease Modeling Meta Regression 2.1 (version 2.1) was used to obtain prepayment and confidence data by location, year, age, and sex for MS. Meanwhile, for areas without data, DisMod MR 2.1 can also use location level covariates to estimate missing data. For the estimation of ASR, GBD 2021 used the GBD standard population structure for multiple calculations, and the final estimate is the average estimate. In previous versions, GBD performed 1000 calculations, but in GBD 2021, only 500 calculations were performed because it was found in simulation testing that the final estimate was not affected by this reduction. ASR data from different countries and regions can be compared across regions due to the use of the same GBD standard population structure. At the same time, GBD 2021 also provides a 95% uncertainty interval for ASR, which provides a more reasonable indicator for our disease burden research.^2^

In order to study the disease burden of MS more comprehensive, we extracted the number, percent and rate of incidence, prevalence, deaths and DALY data of MS in the GBD database, and studied 204 countries, 21 regions, and 5 SDI regions in the world. For analyze the influence of age and gender on MS, we also extracted 10 age groups spanning 1 to 99 years old and gender data. The data encompasses the years 1990-2021 and includes the annual rate of change. Smoking is the only environmental risk among the 84 risks quantified in GBD 2021, and it already has sufficient evidence for a causal relationship with multiple sclerosis. In addition, we obtained the percentage of DALYs for MS due to smoking risk factors versus the number of deaths reported in GBD 2021.^3^

#### Confidence interval and uncertainty interval

Uncertainty intervals (UI) and confidence intervals (CI) serve as statistical measures to gauge the potential range of a population parameter, reflecting the degree of certainty associated with an estimate. These intervals encompass any quantification of estimation uncertainty, not necessarily tied to a predefined confidence level. They are employed to illustrate the variability in estimates due to factors such as measurement error, sampling variability, or inherent population diversity. While UI is not inherently linked to a probability, they provide insight into the possible range of the true parameter. There is no conceptual difference between UI and CI, often associated with a designated confidence level, such as 95%, and these represent the range within which the true population parameter is expected to lie with that probability. Confidence intervals are frequently used in hypothesis testing and data analysis to assess the reliability of an estimate, and they mathematically mirror the estimation of sample statistic uncertainty.^4^

#### Socio-demographic Index

The Socio-demographic Index (SDI), a composite measure, quantifies a country's development level by integrating three fundamental components: lag-distributed income per capita, average years of education for individuals aged 15 and above, and the total fertility rate (TFR) for females under 25. Each component undergoes a rescaling process using specific health-related benchmarks to attain a value within the range of 0·005 to 1. The SDI is subsequently derived as the geometric mean of these adjusted values, multiplied by 100. This index serves as an indicator of the underlying social and economic factors that impact health outcomes within a given region, with higher scores indicating a more advanced socioeconomic status. The Global Burden of Disease (GBD) 2021 study's 204 countries and territories are subsequently classified into five distinct SDI categories: low, low-middle, middle, high-middle, and high.^3^

#### Mendelian randomization(MR) and principal component analysis(PCA)

MR is an analytical method that utilizes genetic variations strongly associated with exposure or risk factors as instrumental variables to evaluate the causal relationship between exposure factors and clinically relevant outcomes.^5^ The MR Analysis includes two parts: exploring the relationship between MS and smoking, and searching for new risk factors. For the first parts: genome-wide association study (GWAS) summary data for MS were obtained from ieu-a-821 of the Integrative Epidemiology Unit open GWAS project (Open GWAS, https://gwas.mrcieu.ac.uk) study and included 978 multiple sclerosis (MS) cases from the European region .^6^ The GWAS summary data on smoking contains 424,960 smoking patients from Europe with the ID number ukb-b-2134. For the second parts: GWAS summary data for MS were obtained from ieu-b-18 and included 47429 MS cases from the European region.^7^ GWAS summary data for BMI came from ieu-a-2 and included 339224 sample.^8^ GWAS summary data for waist circumference came from ieu-a-65 and included 104405 sample.^9^ The data for HDL-C were obtained from ieu-a-780 and included 92860 sample from the European region.^10^ Overweight came from ieu-a-93, which had 158855 sample.^11^

The screening criteria for instrumental variables (IV) are as follows: (1) single nucleotide polymorphism (SNP) associated with smoking with a P-value less than 5e-8 in GWAS are instrumental variables. (2) Using the European 1000 Genomes Project sample as a reference panel, the linkage disequilibrium (LD) among SNPs was calculated with a window value of 5,000 and an R² of 0·1. (3) The intersection of SNPs of MS and risk factors. Next, the R package "TwoSampleMR" was used to determine the relationship between MS and smoking.

PCA uses the same body mass index, waist circumference, and overweight data as MR analysis. Filter through the "mv_detracte_exposures" function in the R package "TwoSampleMR", with the same filtering conditions of a window value of 5000 and an R² of 0.1 as MR analysis.To ensure a strong correlation between instrumental variables and exposure, the p-values of all SNPs are less than 5e-8. Then use the "mv_ramonise_data" function to extract the overlapping SNPs between MS and these three risk factors. Afterwards, use the R package "psych" to perform PCA analysis on the beta values of these three risk factors.

### Result

This section complements the results in the main text by providing more detailed results for the four indicators at the global, regional and country levels. In addition, the percentage change in MS deaths due to smoking and the MR results for smoking and MS are shown here.

#### The global burden of MS from 1990 to 2021

*Prevalence*

From 1990 to 2021, the global prevalence of MS was 1,004,659 (95% uncertainty interval (UI): 868,374 to 1,165,224) cases, and by 2021, this number had increased to 1,887,767 (95% UI: 1,688,654 to 2,113,707) cases, representing 87·901% (95% UI: 80·847% to 95·534%) increase compared to 1990. The global ASPR from 22·26 (95% UI: 19·3 to 25·65) to 22·17 (95% UI: 19·77 to 24·82) with an AAPC was -0·0108 (95% CI: -0·0348 to -0·0132) from 1990 to 2021 (Table S1).

*Incidence*

In 1990, there were 41,970 (95% UI: 36,605 to 48,234) incidence cases of MS around the world, and the number of incidence cases in 2021 was 62,920 (95% UI: 56,015 to 70,635), with an increase of 49·916% (95% UI: 44·654% to 55·226%) compared with 1990. The global ASIR declined from 0·8 (95% UI: 0·7 to 0·91) in 1990 to 0·78 (95% UI: 0·69 to 0·87) in 2021 with an AAPC of -0·1126 (95% CI: -0·1317 to -0·0934) from 1990 to 2021 (Table S1).

*DALYs*

At the global level, there were a total of 574,234 (95% UI: 496,155 to 662,160) DALYs attributed to MS in 1990, which rose to 973,298 (95% UI: 838,209 to 1,133,291) DALYs in 2021, indicating a rise of 69·495% (95% UI: 63·584% to 75·566%). The ASDR was 12·78 (95% UI: 11·1 to 14·72) in 1990 and declined to 11·37 (95% UI: 9·77 to 13·23) in 2021 with the AAPC from 1990 to 2021 was -0·3713 (95% CI: -0·4928 to -0·2496) (Table S1).

*Deaths*

From 1990 to 2021, the deaths due to MS globally from 9,107 (95% UI: 8,711 to 9,469) increased to 16,302 (95% UI: 15,357 to 17,039) representing a 78·992% (95% UI: 68·957% to 88·431%) increase over the period. The global ASMR was 0·22 (95% UI: 0·21 to 0·23) declined to 0·19 (95% UI: 0·18 to 0·2) and the AAPC was -0·4437 (95% CI: -0·6408 to -0·2462). (Table S1)

#### The regional burden from 1990 to 2021

*Prevalence*

At the regional level, the top three regions with the highest ASPRs in 2021 were High-income North America (103·61, 95% UI: 96·44 to 111·33), Western Europe (91·36, 95% UI: 80·9 to 102·99), and Australasia (59·95, 95% UI: 51·99 to 68·82). In contrast, the top three regions with the lowest ASPRs were Oceania (1·6, 95% UI: 1·2 to 2·1), East Asia (2·31, 95% UI: 1·81 to 2·92), and Southeast Asia (2·4, 95% UI: 1·87 to 3·04) (Table S1).

Between 1990 and 2021, the prevalence of cases increased in all regions across the globe. The region with the largest percentage change is Central Latin America (262·506%, 95% UI: 236·561% to 289·627%), followed by Andean Latin America (256·159%, 95% UI: 234·809% to 277·522%), as well as North Africa and Middle East (233·915%, 95% UI: 223·826% to 244·953%). It is noteworthy that Central Europe (18·928%, 95% UI: 14·239% to 24·897%) and Eastern Europe (18·599%, 95% UI: 9·325% to 30·644%) have experienced the smallest increases, which are significantly lower than those observed in other regions (Table S1).

From 1990 to 2021, the largest increase in ASPR of MS was observed in Central Latin America (AAPC: 1·4686, 95% CI: 1·4402 to 1·497), followed by Australasia (AAPC: 1·3172, 95% CI: 1·1703 to 1·4643) and Andean Latin America (AAPC: 1·2519, 95% CI: 1·2385 to 1·2653). The data indicates that ASPR decreased solely in Central Asia (AAPC: -0·03, 95% CI: -0·0621 to -0·0022), while it increased in all other regions (Table S1).

*Incidence*

The top three regions with the highest ASIRs in 2021 were also High-income North America (3·58, 95% UI: 3·33 to 3·86), Western Europe (3·58, 95% UI: 2·93 to 3·66), and Australasia (2·19, 95% UI: 1·92 to 2·47). Whereas, Oceania (0·15, 95% UI: 0·12 to 0·19), East Asia (0·17, 95% UI: 0·14 to 0·2), and Southeast Asia (0·19, 95% UI: 0·15 to 0·22) showed the lowest rates (Table S1).

Between 1990 and 2021, the incidence cases rose in all global regions with the exception of Eastern Europe (-27·578%, 95% UI: -32·03% to -22·38%) and Central Europe (-22·317%, 95% UI: -25·13% to 18·997%). Western Sub-Saharan Africa (213·837%, 95% UI: 202·22% to 228·34%), Andean Latin America (182·053%, 95% UI: 162·544% to 201·134%), and Central Sub-Saharan Africa (174·215%, 95% UI: 163·753% to 185·035%) were the top three regions with the largest increases (Table S1).

From 1990 to 2021, the top three regions with largest increase in ASIR were Central Latin America (AAPC: 1·0944, 95% CI: 1·0717 to 1·1172), Australasia (AAPC: 1·0868, 95% CI: 1·0012 to 1·1725), and Andean Latin America (AAPC: 0·8912, 95% CI: 0·8798 to 0·9027). The regions of Eastern Europe (AAPC: -0·3558, 95% CI: -0·373 to -0·3386) and Central Asia (AAPC: -0·3166, 95% CI: -0·3647 to -0·2686) experienced the largest declines in ASIR, with the decline significantly surpassing that of other regions (Table S1).

*DALYs*

High-income North America (49·15, 95% UI: 41·84 to 56·53), Western Europe (45·33, 95% UI: 38·18 to 52·08), and Central Europe (29·85, 95% UI: 26·3 to 33·74) were the top three regions with the highest ASDR. In contrast, the region with the lowest ASDR was Oceania (0·46, 95% UI: 0·3 to 0·66), followed by East Asia (0·86, 95% UI: 0·63 to 1·13) and Southeast Asia (1·18, 95% UI: 0·94 to 1·45) (Table S1).

Between 1990 and 2021, DALYs increased in all regions of the world with the exception of Eastern Europe (-23·803%, 95% UI: -31·157% to -16·377%) and Central Europe (-21·161%, 95% UI: -27·326% to -14·895%). Central Latin America (337·257%, 95% UI: 293·212% to 382·961%) was the region with the largest percentage change in DALYs, followed by Western Sub-Saharan Africa (315·185%, 95% UI: 189·871% to 508·26%) and Andean Latin America (294·801%, 95% UI: 235·77% to 364·01%) (Table S1).

The top three increasing trends in ASDR belonged to Central Latin America (AAPC: 2·0793, 95% CI: 1·8105 to 2·3488), Andean Latin America (AAPC: 1·5328, 95% CI: 1·1647 to 1·9023) and Western Sub-Saharan Africa (AAPC: 1·3392, 95% CI: 1·2191 to 1·4595). Whereas, Central Europe (AAPC: -1·1941, 95% CI: -1·2980 to -1·0902) showed the largest decreasing trend, followed by Eastern Europe (AAPC: -1·0346, 95% CI: -1·5731 to -0·4931) and Southern Latin America (AAPC: -0·9111, 95% CI: -1·1795 to -0·6419). (Table S1)

*Deaths*

High-income North America (0·81, 95% UI: 0·76 to 0·86), Western Europe (0·73, 95% UI: 0·68 to 0·77), and Central Europe (0·55, 95% UI: 0·5 to 0·61) were the top three regions with the highest ASMR. In contrast, the ASMR of Central Sub-Saharan Africa, East Asia, Eastern Sub-Saharan Africa, High-income Asia Pacific, Oceania, South Asia, and Southeast Asia demonstrated a good performance with all close to 0 (Table S1).

Between 1990 and 2021, change in counts of deaths increased in all regions of the world with the exception of Eastern Europe (-28·448%, 95% UI: -36·808% to -19·298%) and Central Europe (-20·031%, 95% UI: -28·756% to -11·077%). Central Latin America (437·025%, 95% UI: 369·464% to 512·266%) experienced the largest increase in deaths, followed by Andean Latin America (388·025%, 95% UI: 267·406% to 540·825%), as well as North Africa and Middle East (367·814%, 95% UI: 226·187% to 699·212%) (Table S1).

The top three increasing trends in ASMR belonged to Central Latin America (AAPC: 2·2932, 95% CI: 1·8102 to 2·7786), Andean Latin America (AAPC: 2·0478, 95% CI: 1·3576 to 2·7427), and South Asia (AAPC: 2·0361, 95% CI: 1·9191 to 2·1533). Whereas, the three regions experiencing the most significant decline in ASMR, such as Southern Latin America (AAPC: -1·5618, 95% CI: -2·0747 to -1·0462), Central Europe (AAPC: -1·5068, 95% CI: -1·6548 to -1·3586), and Central Asia (AAPC: -1·4921, 95% CI: -2·3928 to -0·583), also exhibited a pronounced downward trend in ASMR (Table S1).

#### The national burden from 1990 to 2021

*Prevalence*

In 2021, ASPR of MS ranged from 1·45 to 161·6 cases per 100,000 population in 204 countries and territories worldwide. Sweden (161·6, 95% UI: 140·22 to 186·95), Canada (134·2, 95% UI: 130·83 to 137·7), and Norway (131·53, 95% UI: 111·04 to 154·17) had the three highest ASPR. Whereas, Papua New Guinea (1·45, 95% UI: 1·08 to 1·91), Nauru (1·56, 95% UI: 1·18 to 1·99), and Kiribati (1·74, 95% UI: 1·33 to 2·22) showed the lowest rate (Figure 1, Table S2).

The increase of the relative change in the prevalence cases of MS between 1990 and 2021 was most noticeable in Qatar (1384·979%, 95% UI: 1184·123% to 1626·707%), followed by United Arab Emirates (871·967%, 95% UI: 778·484% to 1004·875%) and Kuwait (623·733%, 95% UI: 571·261% to 681·832%). It is worth noting that all three countries are from the Middle East region. In 2021, only nine countries globally reported fewer cases than in 1990, these countries were Bulgaria, Estonia, Hungary, Latvia, Lithuania, Niue, Romania, Ukraine, as well as Bosnia and Herzegovina, which are predominantly located at distance from the equator. The three countries with the highest decrease were Hungary (-20·79%, 95% UI: -26·673% to -14·359%), Latvia (-11·206%, 95% UI: -16·636% to -5·113%), and Ukraine (-4·435%, 95% UI: -10·841% to -1·948%) (Figure S1, Table S2).

From 1990 to 2021, the two countries with the highest AAPC of ASPR were Ghana (1·9143, 95% CI: 1·8713 to 1·9574) and Egypt (2·5161, 95% CI: 2·3472 to 2·6852). Hungary (-0·7442, 95% CI: -0·7921 to -0·6962) and Uzbekistan (-0·7167, 95% CI: -0·7584 to -0·6751) had the most obvious decreasing trends in ASPR (Figure S2, Table S2).

*Incidence*

In 2021, ASIR varied from 0·1 to 5·6 cases per 100,000 population in 204 countries and territories worldwide. Sweden (5·58, 95% UI: 4·91 to 6·34), Norway (4·87, 95% UI: 4·18 to 5·61), and Canada (4·74, 95% UI: 4·64 to 4·86) were the three countries with the highest ASIRs in the world. Whereas, the ASIRs of Papua New Guinea, Maldives, Guam, and Malaysia were 0·15 (95% UI: 0·11 to 0·18), 0·13 (95% UI: 0·11 to 0·16), 0·15 (95% UI: 0·12 to 0·18), and 0·15 (95% UI: 0·12 to 0·18), making them the lowest ASIR countries in the world (Figure S3, Table S3).

The magnitude of increase of the relative change in the incidence cases of MS between 1990 and 2021 was also most noticeable in Qatar (1151·084%, 95% UI: 979·237% to 1360·543%), followed by United Arab Emirates (502·734%, 95% UI: 389·972% to 629·651%) and Oman (400·487%, 95% UI: 340·922% to 455·668%). Similar to the magnitude of increases in prevalence cases, Qatar and United Arab Emirates also had the highest increases in incidence. In contrast, there were 22 countries or regions in the world with a decrease in the number of incidence cases, and the three with the largest decrease were Latvia (-40·455%, 95% UI: -43·771% to -36·568%), Hungary (-39·936%, 95% UI: -43·459% to -35·913%), and Lithuania (-37·089%, 95% UI: -40·43% to -33·206%) (Figure S4, Table S3).

From 1990 to 2021, Egypt (AAPC: 2·0768, 95% CI: 1·9119 to 2·2421), Kuwait (AAPC: 1·6329, 95% CI: 1·6047 to 1·6612), and Ghana (AAPC: 1·5991, 95% CI: 1·5505 to 1·6478) had the highest increasing trend in ASIR. Whereas, Hungary (AAPC: -0·9622, 95% CI: -1·0717 to -0·8526), Uzbekistan (AAPC: -0·9392, 95% CI: -0·971 to -0·9074), and Albania (AAPC: -0·7226, 95% CI: -0·7568 to -0·6884) showed the highest decreasing trend among 57 locations of decline (Figure S5, Table S3).

*DALYs*

The top three countries with the highest ASDR in 2021 were United Kingdom (71·3, 95% UI: 61·83 to 80·79), Denmark (65·78, 95% UI: 56·6 to 76·05), and Sweden (63·51, 95% UI: 51·03 to 77·77). Whereas, Nauru and Papua New Guinea showed the lowest ASDR at 0·44 (95% UI: 0·29 to 0·64) and 0·41 (95% UI: 0·27 to 0·6). (Figure S6, Table S4)

The most noticeable increase of DALYs was observed in Qatar (1,380·277%, 95% UI: 1,094·6% to 1,778·844%), followed by United Arab Emirates (831·433%, 95% UI: 639·928% to 1,074·662%) and Kuwait (686·908%, 95% UI: 564·158% to 852·038%). Only 16 locations worldwide experienced a decrease in DALYs, and the three with the largest decrease were Estonia (-61·018%, 95% UI: -67·916% to -54·193%), Latvia (-57·532%, 95% UI: -64·649% to -50·183%), and Lithuania (-49·696%, 95% UI: -58·325% to -40·594%) (Figure S7, Table S4).

From 1990 to 2021, Mauritius (AAPC: 4·6177, 95% CI: 3·7486 to 5·4941) had the largest increase in ASDR, much more than other locations, with Libya (AAPC: 2·97, 95% CI: 2·9007 to 3·0393) coming in second. The location where ASDR decrease was similar to where ASDR increase, with the most obvious decreasing trends was Estonia (AAPC: -2·8946, 95% CI: -3·2851 to -2·5026), where the decline was considerably steeper than other locations (Figure S8, Table S4).

*Deaths*

The numbers of ASMR in MS is relatively low in 2021, with the top three countries were United Kingdom (1·34, 95% UI: 1·27 to 1·4), Denmark (1·23, 95% UI: 1·1 to 1·36), and Albania (1·08, 95% UI: 0·59 to 1·83). And 20 other locations like Zimbabwe, Nauru, and Fiji have zero ASMR in 2021 (Figure S9, Table S5).

The most noticeable increase of deaths cases was observed in Kuwait (226,819·954%, 95% UI: 185,058·471% to 279,906·111%), followed by Mauritius (90,722·859%, 95% UI:73,594·421% to 109,252·318%) and Bahrain (7,476·68%, 95% UI: 5,133·666 to 10,788·629), far more than anywhere else. And where there 17 locations were decline in cases, the highest decrease was in Estonia (-59·766%, 95% UI: -67·496% to -51·159%) (Figure S10, Table S5).

From 1990 to 2021, Kuwait (AAPC: 18·8762, 95% CI: 13·2837 to 24·7448), followed by Mauritius (AAPC: 20·4691, 95% CI: 18·7182 to 22·2458) and Bahrain (AAPC: 7·8776, 95% CI: 7·2295 to 8·5296) also demonstrated the largest increase in ASMR. Once again Estonia (AAPC: -3·2316, 95% CI: -3·6566 to -2·8048) showed the most obvious decreasing trends (Figure S11, Table S5).

#### The impact of risk Factors on the disease burden

The percentage of deaths due to smoking-related MS was similar to the overall trend in DALY, showing an overall decrease in 2021 compared to 1990. From 1990 to 2021, the global percentage of deaths due to MS caused by smoking decreased from 18% to 11%. Similarly, in the five SDI regions, there was a significant decline in the percentage of the high SDI region (from 21·1% to 12·8%) and, as a result, the high-middle SDI region had the largest percentage in 2021.The largest region in terms of percentage was High-income North America in 1990 and Central Europe in 2021 (23·9% and 14·5% respectively), while the smallest region was Western Sub-Saharan Africa. The difference was that in addition to Eastern Europe (from 13·3% to 14·4%), there was also an remains unchanged in Central Asia (8·1%). The distribution trend of risk factors in other regions was basically the same in 1990 as in 2021. Significant gender differences in the contribution of risk factors to deaths were consistent with DALYs (Figure S11).

#### Results of MR and PCA

To investigate whether there is a causal relationship between smoking and MS, we conducted an MR study. According to the screening criteria of IVs, 19 SNPs meeting the criteria were selected as instrumental variables in the Open GWAS database. The outcome factors were from patients with a history of smoking, and the exposure factors were patients with trait identified as MS. Two-sample Mendelian randomization analysis showed a weak link between smoking and MS risk (OR:0·08). (Figure S15). The results suggest that smoking and MS may not be genetically linked. However, smoking does affect the occurrence of MS, and it makes a certain contribution to the DALYs of MS.

We selected SNPs related to exposure as tools to explore the relationship between exposure. After screening, there are still 94 SNPs strongly correlated with BMI, waist circumference, and overweight. PCA analysis was performed on these 94 SNPs, and the loadings on their first principal component (PC1) were found to be 0.995, 0.982, and 0.988, respectively. This indicates that PC1 can be represented by 0.995 BMI , 0.982 waist circumference , and 0.988 overweight. Meanwhile, the contribution of the first principal component is 97.693%, indicating a high correlation between BMI, waist circumference, and overweight. This indicates that these three risk factors may have a unified underlying trait or indicator expression, and these SNPs may affect these phenotypes through common genetic mechanisms. PC1 can be considered as a 'global obesity index'. Afterwards, we conducted MR analysis using PC1 and MS and found that the results of MR were significant, indicating that the potential traits represented by PC1 are related to the risk of MS, suggesting a causal relationship between obesity and MS.

### **Figure**

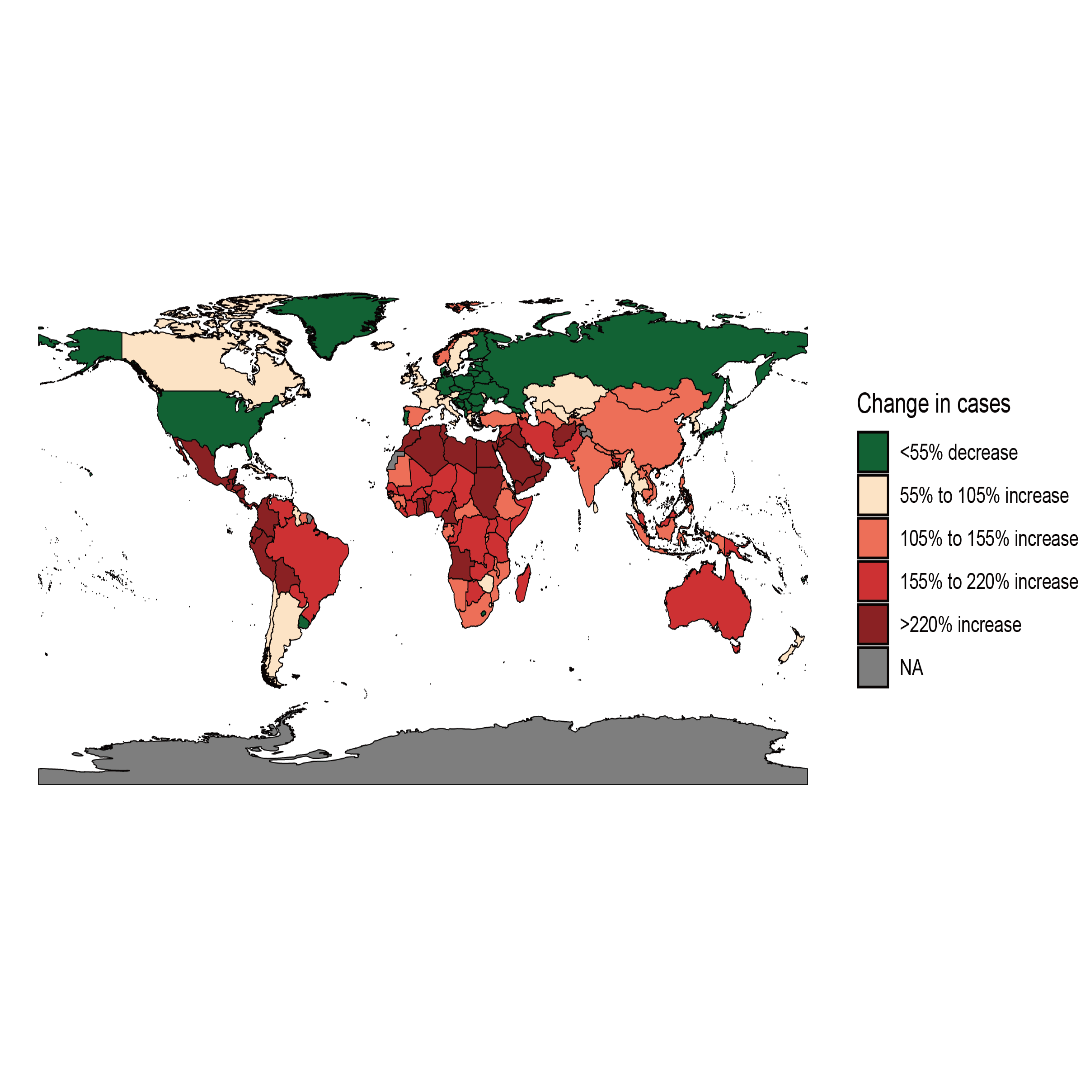

**Figure S1****. World map of the prevalence changing distribution of the number of MS cases from 1990 to 2021.** The black areas represent areas where no or few people live.

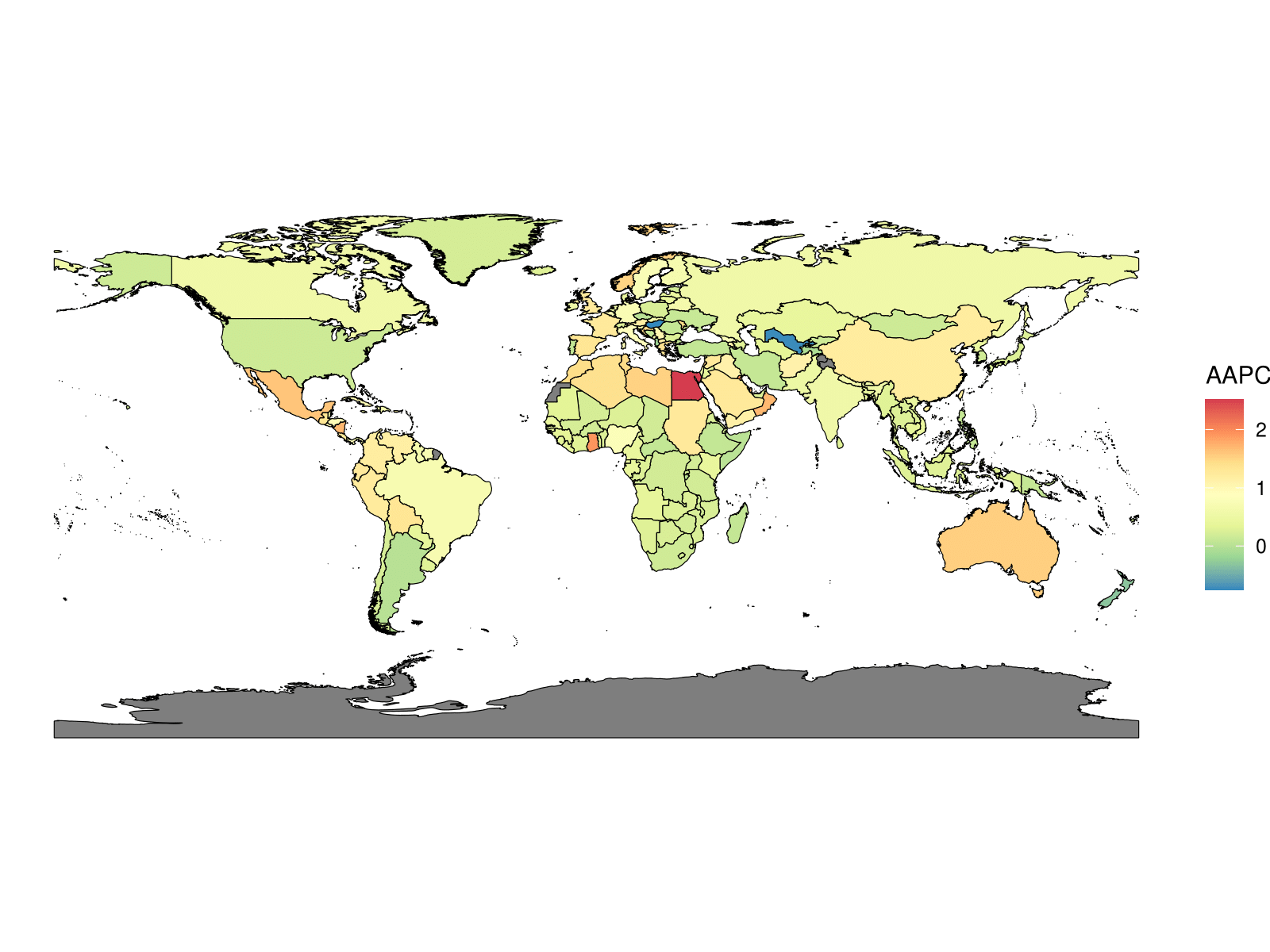

**Figure S2****. MS AAPC of ASPR world map from 1990 to 2021.** The black areas represent areas where no or few people live.

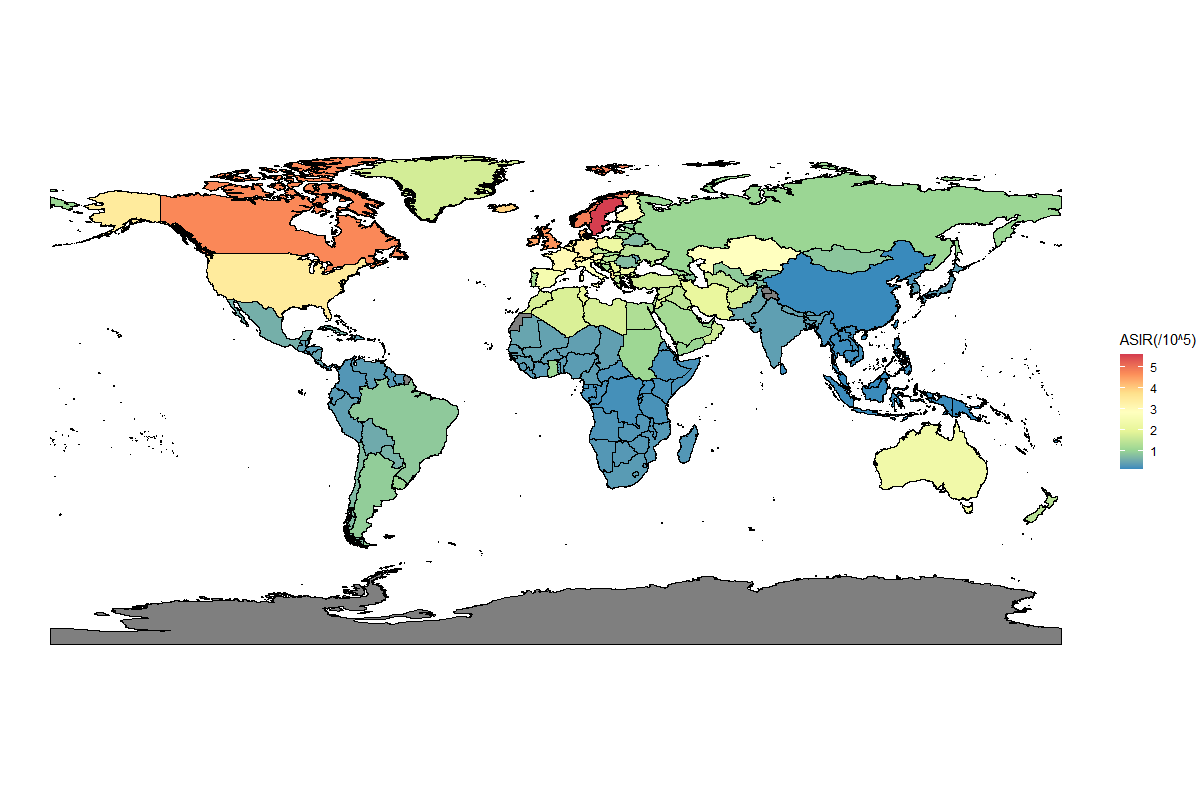

**Figure S3****.World map of ASIR distribution of MS.** The black areas represent areas where no or few people live.

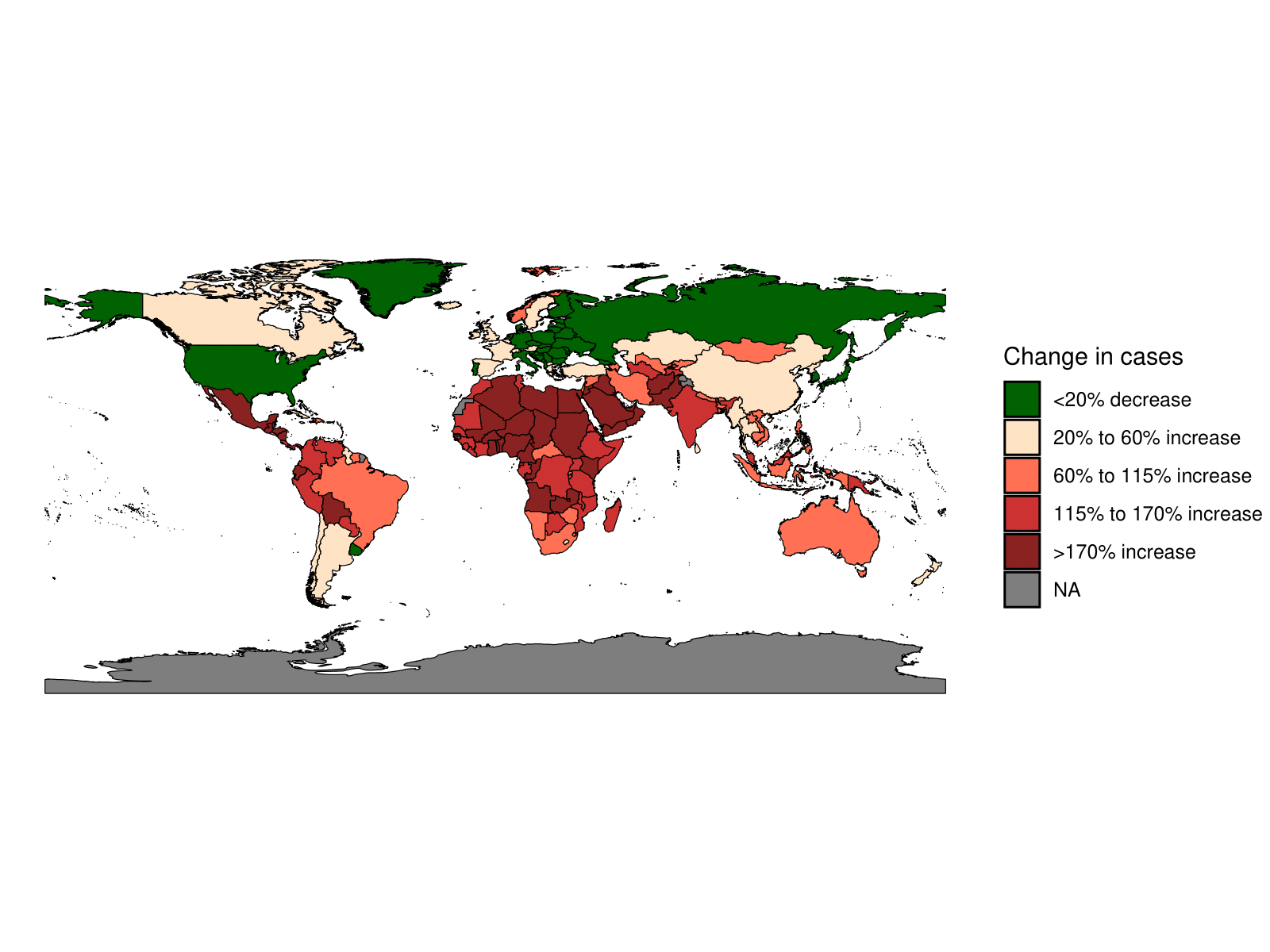

**Figure S4****. World map of the incidence changing distribution of the number of MS cases from 1990 to 2021.** The black areas represent areas where no or few people live.

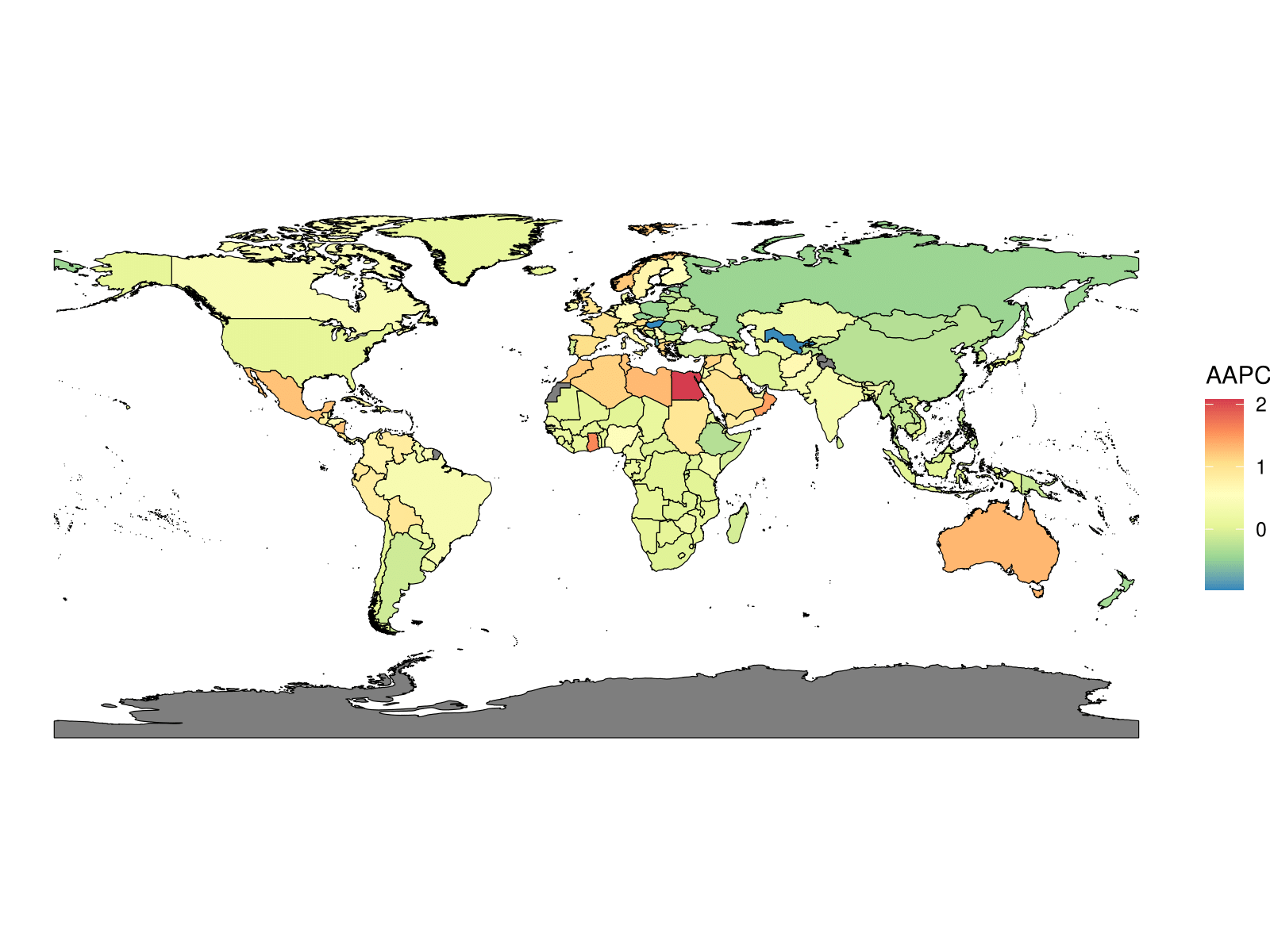

**Figure S5****. MS AAPC of ASIR world map from 1990 to 2019.** The black areas represent areas where no or few people live.

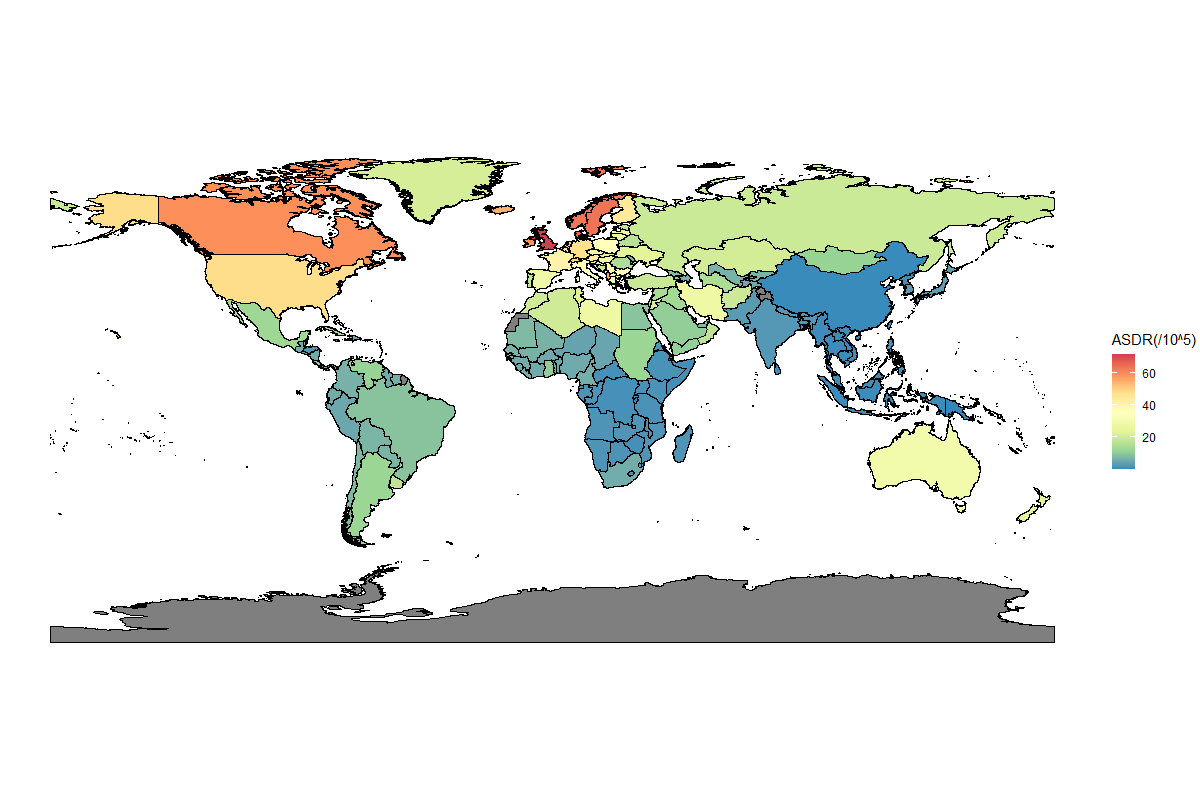

**Figure S6****. World map of ASDR distribution of MS.** The black areas represent areas where no or few people live.

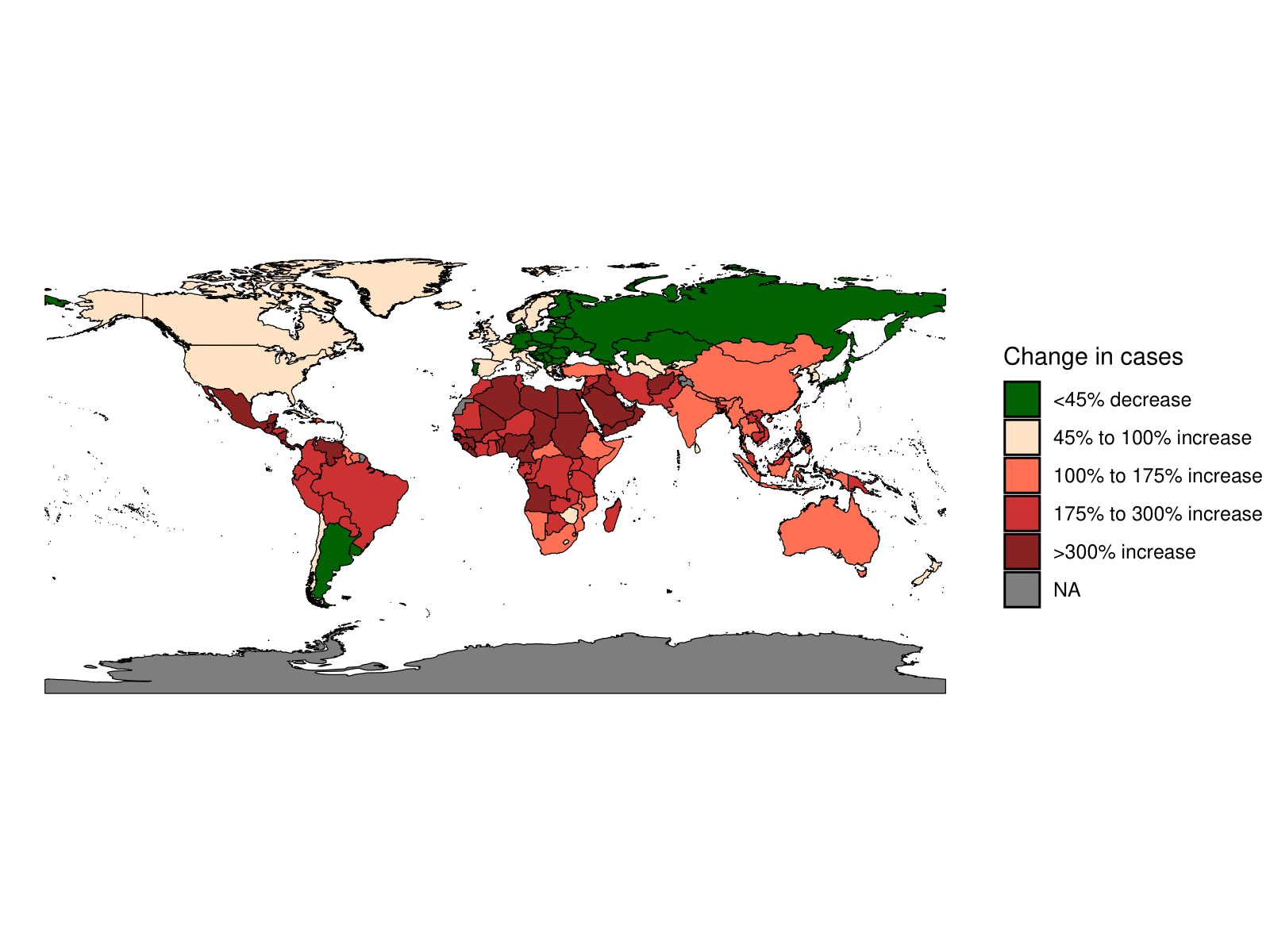

**Figure S7****. World map of the DALYs changing distribution of the number of MS cases from 1990 to 2021.** The black areas represent areas where no or few people live.

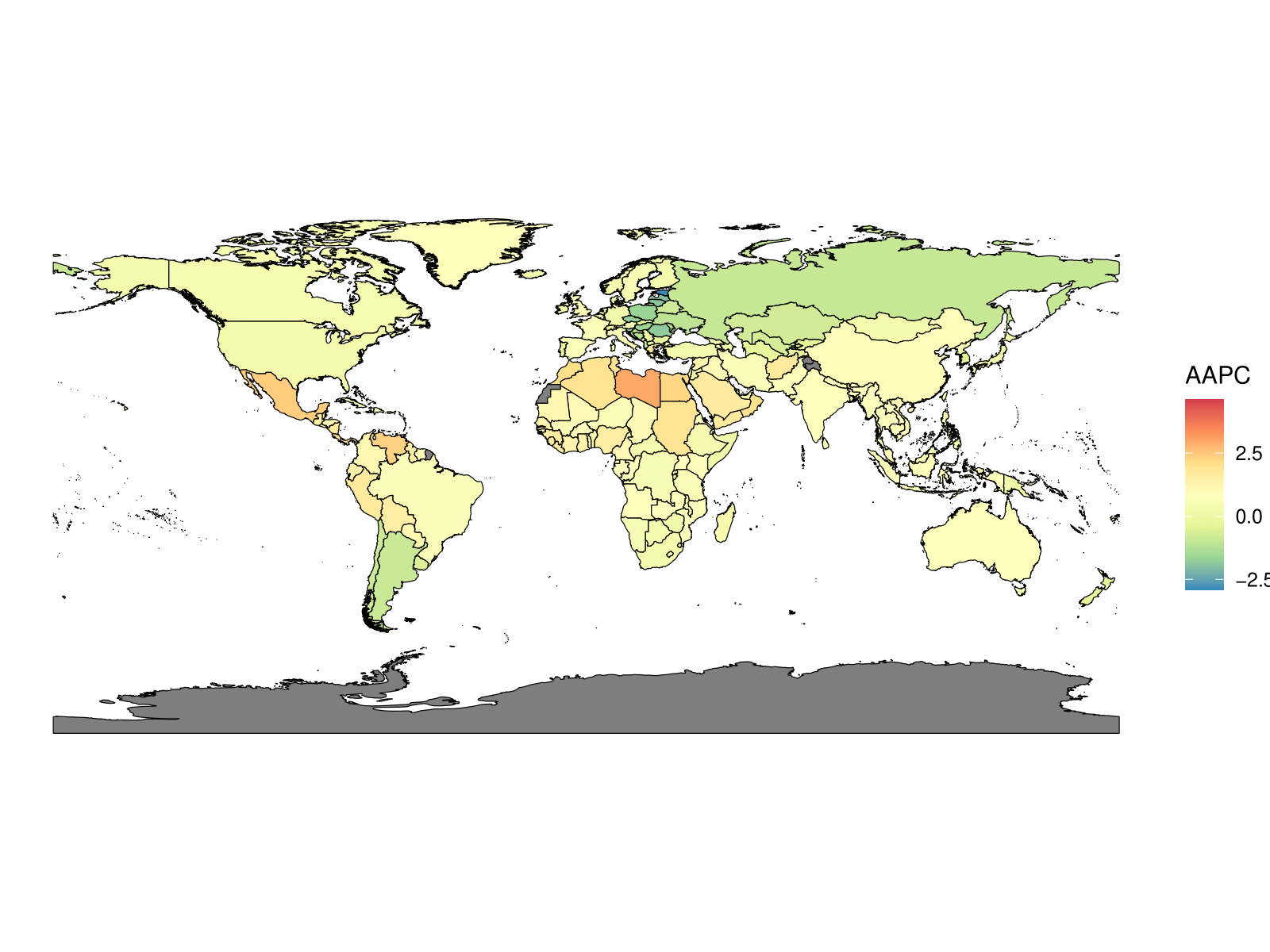

**Figure S8****. MS AAPC of ASDR world map from 1990 to 2021.** The black areas represent areas where no or few people live.

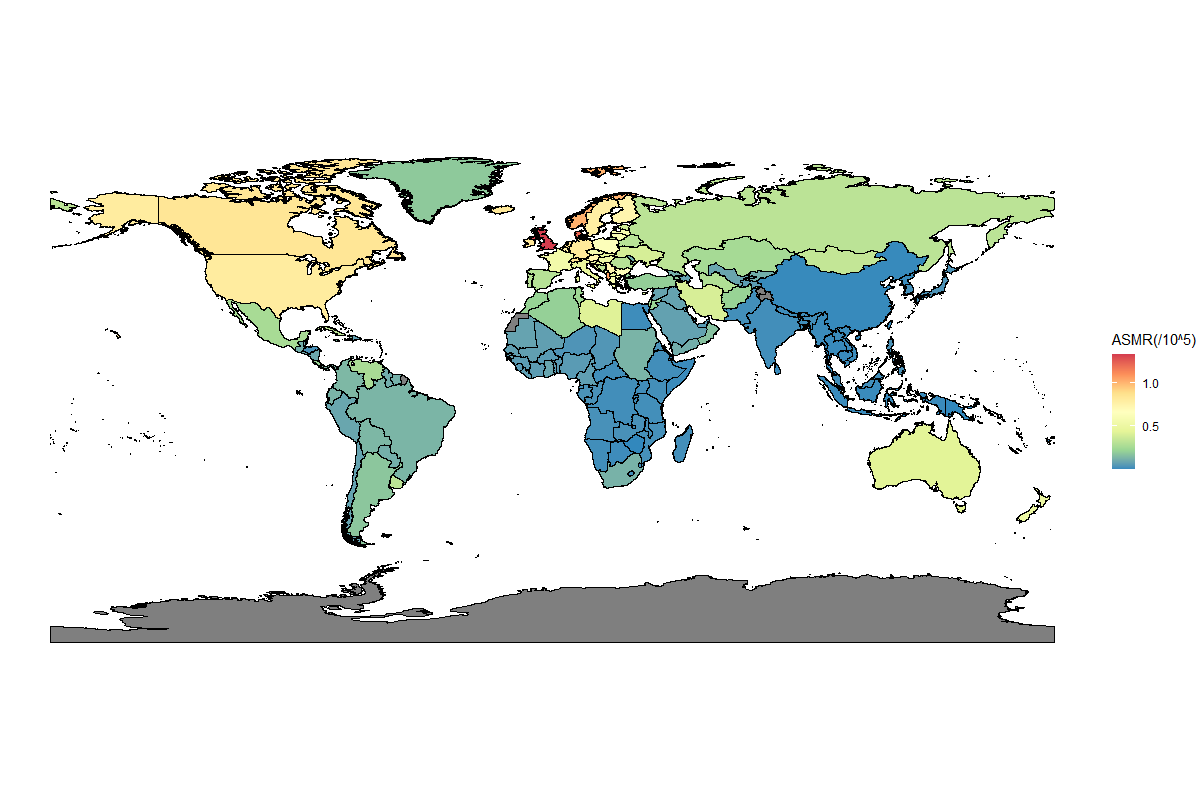

**Figure S9****. World map of ASMR distribution of MS.** The black areas represent areas where no or few people live.

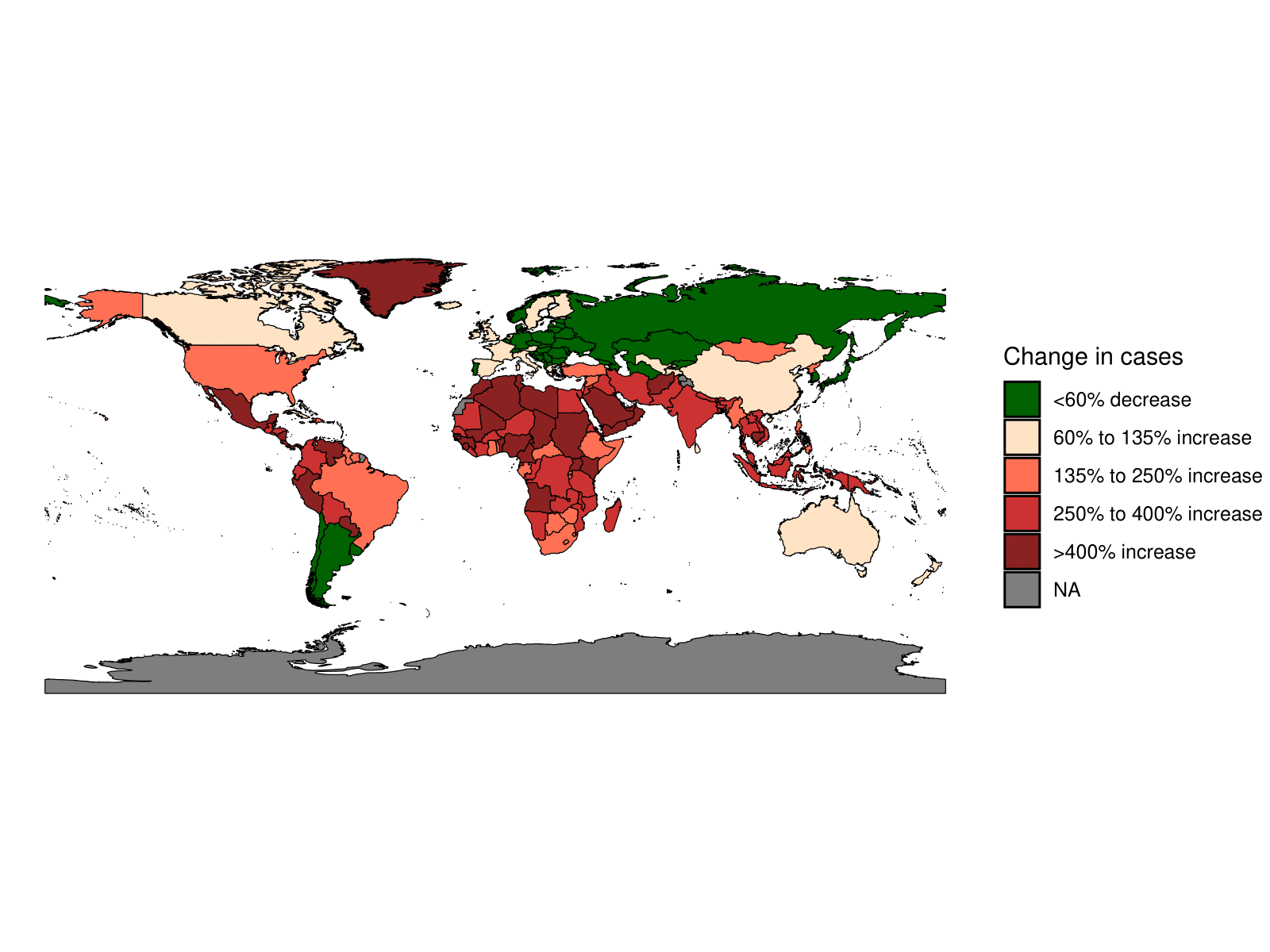

**Figure S10****. World map of the deaths changing distribution of the number of MS cases from 1990 to 2021.** The black areas represent areas where no or few people live.

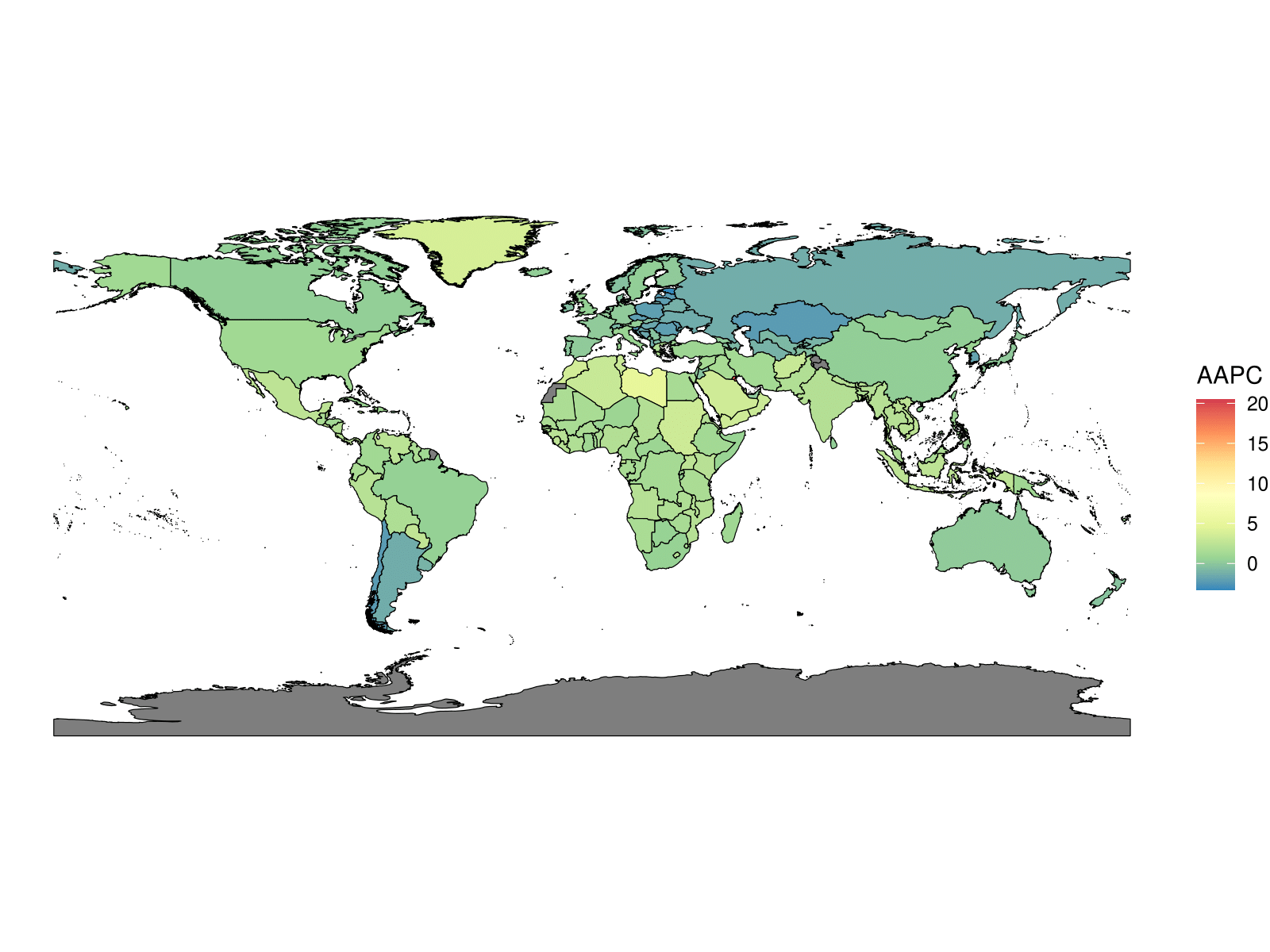

**Figure S11****. MS AAPC of ASMR world map from 1990 to 2021.** The black areas represent areas where no or few people live.

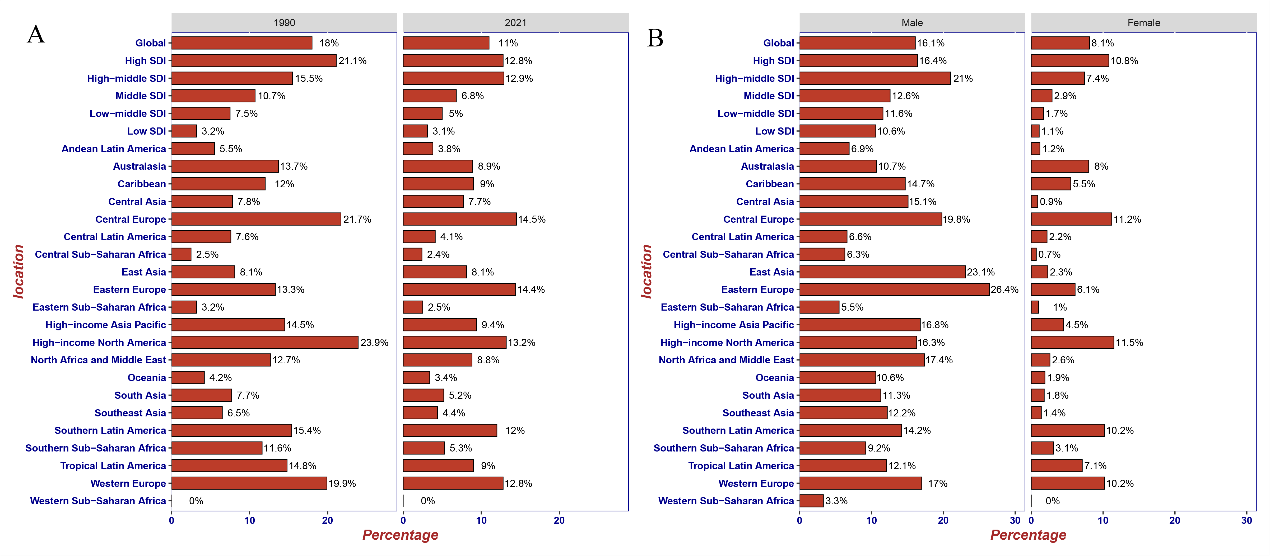

**Figure S12****. Percentage contribution of smoking to MS deaths in 1990 versus 2021 (A) and in male versus female (B) from different GBD regions.**

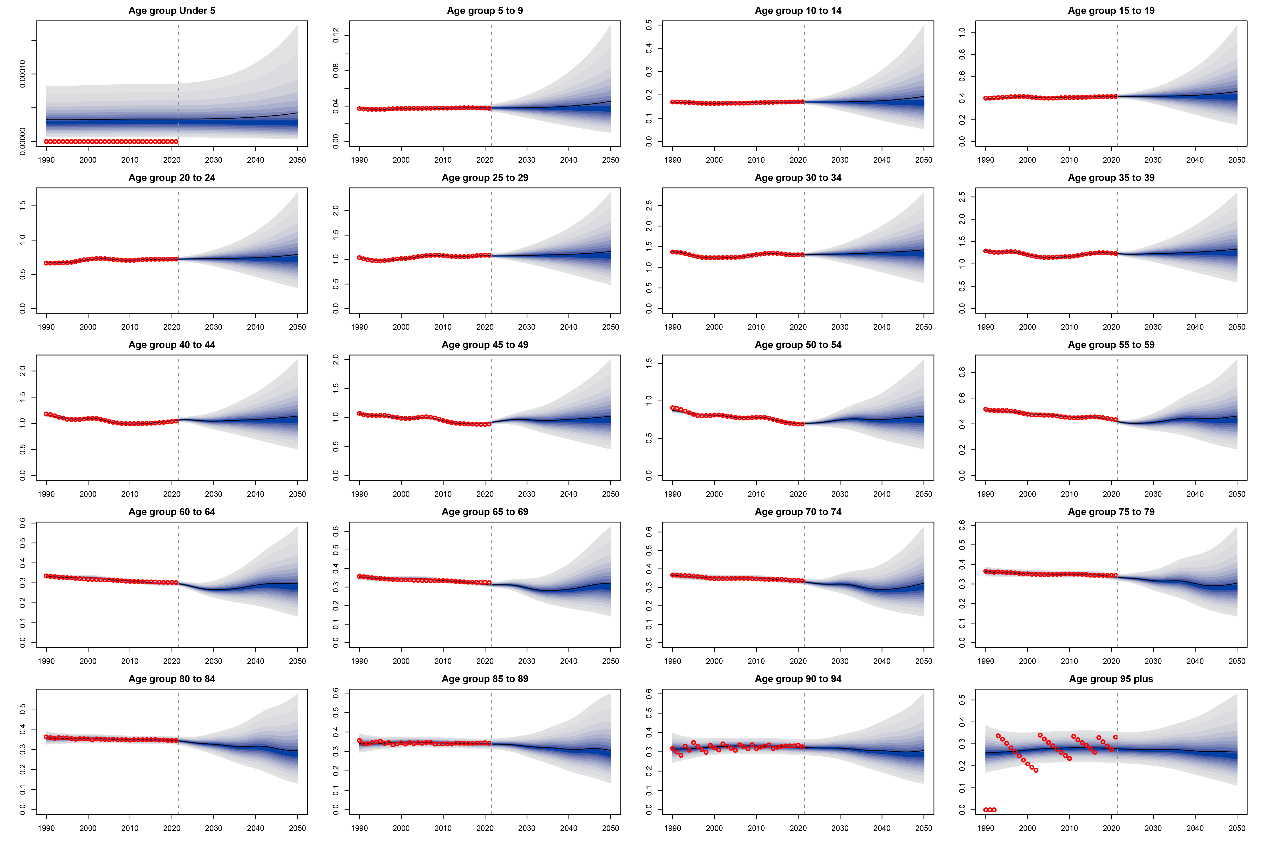

**Figure S13****. Prediction of ASIR development of MS in 10 age groups in male until 2050.**

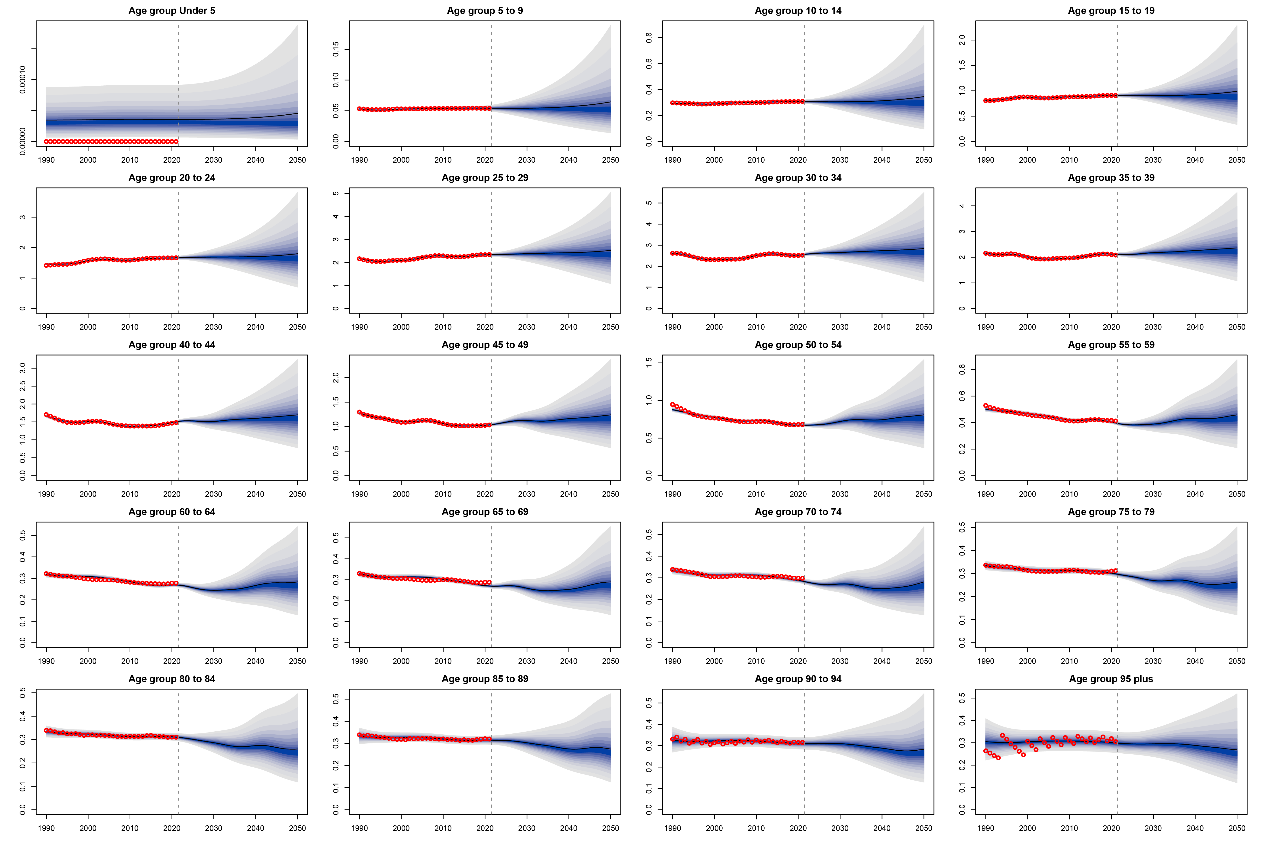

**Figure S14****. Prediction of ASIR development of MS in 10 age groups in female until 2050.**

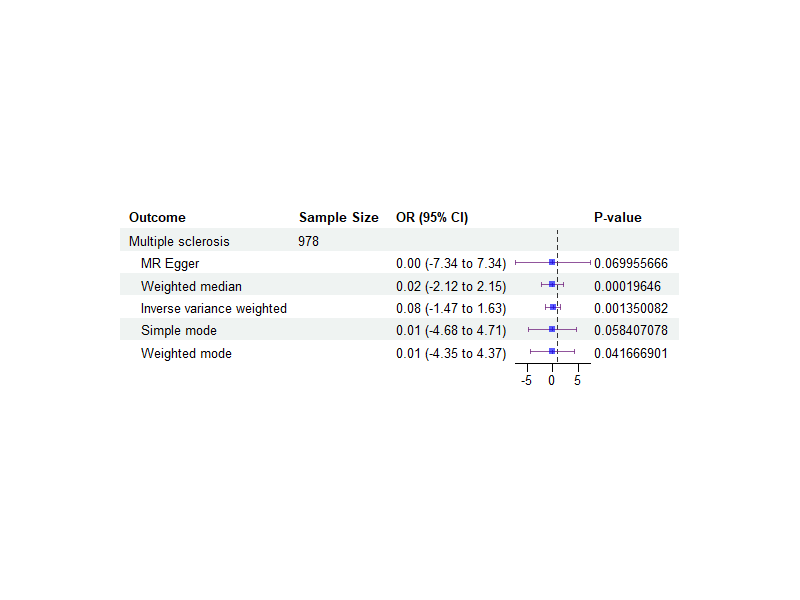

**Figure S15****. MR forest map of smoking on MS.** OR = odds ratio

6.	Baranzini SE, Wang J, Gibson RA, et al. Genome-wide association analysis of susceptibility and clinical phenot ype in multiple sclerosis. *Human molecular genetics*; **18**(4): 767-78.

7.	International Multiple Sclerosis Genetics C. Multiple sclerosis genomic map implicates peripheral immune cells and microglia in susceptibility. *Science (New York, NY)*; **365**(6460): eaav7188.

8.	Locke AE, Kahali B, Berndt SI, et al. Genetic studies of body mass index yield new insights for obesity biol ogy. *Nature*; **518**(7538): 197-206.

9.	Shungin D, Winkler TW, Croteau-Chonka DC, et al. New genetic loci link adipose and insulin biology to body fat distribu tion. *Nature*; **518**(7538): 187-96.

10.	Willer CJ, Schmidt EM, Sengupta S, et al. Discovery and refinement of loci associated with lipid levels. *Nature genetics*; **45**(11): 1274-83.

11.	Berndt SI, Gustafsson S, Mägi R, et al. Genome-wide meta-analysis identifies 11 new loci for anthropometric tr aits and provides insights into genetic architecture. *Nature genetics*; **45**(5): 501-12.
